# A population framework for predicting the proportion of people infected by the far-field airborne transmission of SARS-CoV-2 indoors

**DOI:** 10.1101/2021.11.24.21266807

**Authors:** Christopher Iddon, Benjamin Jones, Patrick Sharpe, Muge Cevik, Shaun Fitzgerald

## Abstract

The number of occupants in a space influences the risk of far-field airborne transmission of SARS-CoV-2 because the likelihood of having infectious and susceptible people both correlate with the number of occupants. This paper explores the relationship between occupancy and the probability of infection, and how this affects an individual person and a population of people. Mass-balance and dose-response models determine far-field transmission risks for an individual person and a population of people after sub-dividing a large *reference* space into 10 identical *comparator* spaces.

For a single infected person, the dose received by an individual person in the *comparator* space is 10-times higher because the equivalent ventilation rate per infected person is lower when the *per capita* ventilation rate is preserved.

However, accounting for population dispersion, such as the community prevalence of the virus, the probability of an infected person being present and uncertainty in their viral load, shows the transmission probability increases with occupancy and the *reference* space has a higher transmission risk. Also, far-field transmission is likely to be a rare event that requires a high emission rate, and there are a set of Goldilocks conditions that are *just right* when ventilation is effective at mitigating against transmission. These conditions depend on the viral load, because when they are very high or low, ventilation has little effect on transmission risk.

Nevertheless, resilient buildings should deliver the equivalent ventilation rate required by standards as minimum.

## 1. Introduction

Severe Acute Respiratory Syndrome Coronavirus 2 (SARS-CoV-2) is a virus that causes COVID-19. In 2020, it spread rapidly worldwide causing a pandemic. The primary mode transmission of the virus occurs when it is encapsulated within respiratory droplets and aerosols and inhaled by a susceptible person [1]. These are most concentrated in the exhaled puff of an infected person, which includes a continuum of aerosols and droplets of all sizes as a multiphase turbulent gas cloud [2, 3]. The subsequent transport of infectious aerosols from the exhaled puff occurs differently in outdoor and indoor environments. Outside, air movement disrupts the exhaled puff, a prodigious space volume rapidly dilutes it [4], and ultra-violet (UV) light renders the virus biologically non-viable over a short period of time [5]. Inside, the magnitude of air movement is usually insufficient to disrupt the exhaled puff, a finite space volume and lower ventilation rates concentrate aerosols in the air, and there is usually less UV light [6]. Accordingly, transmission of the virus occurs indoors more frequently than outdoors [7, 8], and inhaling the exhaled puff at close contact is more likely to lead to an infective dose than when inhaling indoor air at a distance where the virion laden aerosols are diluted. This is consistent with the epidemiological understanding that SARS-CoV-2 is spread primarily by close contact where it might be possible to smell a person’s *coffee breath* [2, 3, 9, 10, 11]. However, it is still possible for a susceptible person to inhale an infective dose of aerosol borne virus, from shared indoor air, known as *far-field* airborne transmission, and occurs at distances of *>* 2 m from the infected person. Far-field transmission is linked to several super spreading events and is often correlated with poor indoor ventilation, long exposure times, and respiratory activities that increase aerosol and viral emission, such as singing [12, 13, 14].

Previous analyses of far-field infection risk consider the presence of a single infected person. However, the number of occupants in a space influences the risk of airborne transmission because the likelihood of having infectious and susceptible people both scale with the number of occupants. Therefore, it may be advantageous to sub-divide large spaces into a number of identical smaller spaces to reduce the transmission risk. Here, the space volume and ventilation rate per person would be kept constant, and occupants equally divided into smaller groups of people. The impact of this strategy on virus transmission is not obvious. On one hand, the smaller space with lower occupancy reduces the probability of an infected person being present, and also reduces the number of susceptible people who are exposed to infected people. On the other hand, the ventilation rate per infected person is likely to be smaller in the smaller space, increasing the transmission risk for any susceptible people present. Accordingly, this paper explores the relationship between occupancy and the probability of infection, and how this affects an individual person and a population of people. We take a theoretical approach to consider the infection risk for the population of a large space and compare it to the same population distributed in a number of smaller identical spaces.

We first consider the infection risk for a person using an existing analytical model [15] to predict the dose, and the probability that the dose leads to infection, in a big and a small space. We then consider the infection risk for two equal populations distributed evenly in either the big space or a number of smaller spaces, by considering the community infection rate, the viral load, and the probability of infection from a viral dose.

Section 2 outlines the modelling approach and the input data. Section 3 considers the personal risks from sub-division and Section 4 considers the risks for a population. Section 5 discusses factors that affect infection risk and limitation of the work.

## 2. Theoretical approach

An analytical model is used to predict the dose of viral genome copies of an individual person and associated individual and population infection risks of infection.

### 2.1. Dose and infection risk

The mass-balance model of Jones *et al.* [15] is used to predict the number of RNA copies absorbed by the respiratory tract of a person exposed to aerosols in well mixed air over a period of time that is sufficient for the viable virus concentration to reach a steady-state, and then combined with the viable fraction, *v*, to give a dose, *D*.

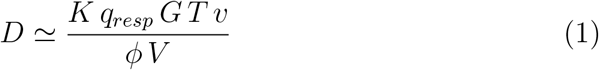

Here, *K* is the fraction of aerosol particles absorbed by respiratory tract, *q_resp_* is the respiratory rate ( m^3^ s*^−^*^1^), *G* is the emission rate of RNA copies ( RNA copies s*^−^*^1^) and is a function of the respiratory activity (see Jones *et al.*), *T* is the exposure period (s), *φ* is the total removal rate ( s*^−^*^1^), which represents the sum of all removal by ventilation, surface deposition, biological decay, respiratory tract absorption, and filtration, and *V* is the space volume (m^3^). The product *φ V* can be considered to be an *equivalent* ventilation rate. The approach is common and has been used by others to investigate exposure in well mixed air [16, 17].

For a full description of the model, a discussion of uncertainty in suitable inputs, and a sensitivity analysis, see Jones *et al.* [15]. The analysis shows that the most sensitive parameter is *G*, the rate of emission of RNA copies. *G* is a function of the *viral load* in the respiratory fluid, *L* ( RNA copies ml*^−^*^1^) and the volume of aerosols emitted, which in turn is a function of exhaled breath rate and respiratory activity; see Appendix A. The distribution of the viral load within the infected population is reported to be log-normal by Yang *et al.* [4], Weibull by Chen *et al.* [18], and Gamma by Ke *et al.* [19]. This suggests that the true distribution is unknown and so we use the data of Chen *et al.* [20] who predict that log_10_ values of viral load are normally distributed with a mean of 7 log_10_ RNA copies ml*^−^*^1^; see Table 2 and Figure 2. We explore variations in these values in Section 2.3 and discuss their origin, and uncertainty in them, in Section 5.5. The probability of a viral load, *P* (*L*), can then be determined from a Gaussian probability density function. The dose can be used to estimate a probability of infection using a dose-response curve. However, there is no dose-response curve for SARS-CoV-2. A number of studies [21, 16, 22] apply a dose curve for the SARS-CoV-1 virus, which is a typical dose curve for corona viruses, and so it is applied here. There are obvious problems with this extrapolation and they are discussed in Section 5.5. The probability of infection of an individual person, *P* (*R*), is assumed to follow a Poisson distribution

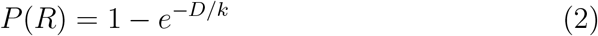

where, *k* is the reciprocal of the probability that a single pathogen initiates an infection. When *D* = *k*, *P* (*R*) = 63%. We use a value of *k* = 410 following DeDiego *et al.*[23].

**Table 1:** General scenario inputs (top) and calculations of individual risk (bottom).

|  | Big Office<br>Reference | Small Office<br>Comparator |
| --- | --- | --- |
| Number of occupants, $N$ | 50 | 5 |
| Space Volume, $V$ ($\text{m}^3$) | 1500 | 150 |
| Air flow rate, $\psi V$ ($\text{ls}^{-1}$) | 500 | 50 |
| Equivalent ventilation rate, $\phi V$ ($\text{ls}^{-1}$) | 942 | 94.2 |
| Air change rate, $\psi$ ($\text{h}^{-1}$) | | 1.2 |
| Removal rate, $\phi$ ($\text{h}^{-1}$) | | 2.26 |
| *Per capita* volume, $V N^{-1}$ ($\text{m}^3$ per person) | | 30 |
| Exposure time, $T$ (h) | | 8 |
| Dose constant [23], $k$ | | 410 |
| Respiratory tract absorption fraction, $K$ | | 0.55 |
| Viable fraction, $v$ (%) | | 100 |
| Respiratory activity, *breathing:talking* (%) |  | 72:25 |
| Volumetric ratio of exhaled droplets to exhaled air, $V_{drop}^*$ | $5.05 \times 10^{-13}$ | |
| Respiratory fluid density ($\text{ml m}^{-3}$) | $1.25 \times 10^8$ | |
| Respiratory rate, $q_{resp}$ ($\text{m}^3 \text{h}^{-1}$) | 0.56 | |
| Viral emission rate, $G$ (RNA copies $\text{h}^{-1}$) | 394 | |
| Community infection rate, $C$ | 1:100 | |
| Viral load [20], $L$ (RNA copies $\text{ml}^{-1}$) | $1 \times 10^7$ | |
| Dose, $D$ (viable virions inhaled) | 0.245 | 2.450 |
| REI | 1 | 10 |

All values converted to SI units before application.

**Table 2:** Scenario inputs (top) and calculations of population risk (bottom) given to 2 significant figures.

|  | Big Office<br>Reference | Small Office<br>Comparator |
| --- | --- | --- |
| Viral load [20] ($\log_{10}$ RNA copies $\text{ml}^{-1}$) | N(7,1.4) | |
| Population, $N_{pop}$ | $1 \times 10^7$ | |
| Number of spaces | $2 \times 10^6$ | $2 \times 10^5$ |
| Probability of transmission event, $P(0 < I < N)$ (%) | 39 | 4.9 |
| Probability of susceptible people, $P(S)$ (%) | 38 | 3.9 |
| Mean number of infected people$^{\ddagger}$, $\bar{I}$ | 1.3 | 1.0 |
| Mean emission rate$^{\dagger}$ (RNA copies $\text{h}^{-1}$) | $2.5 \times 10^4$ | $2.6 \times 10^3$ |
| Mean emission rate$^{\ddagger}$ (RNA copies $\text{h}^{-1}$) | $6.3 \times 10^4$ | $5.4 \times 10^4$ |
| Mean dose$^{\dagger}$ (virions inhaled) | 17 | 18 |
| Mean dose$^{\ddagger}$ (virions inhaled) | 44 | 370 |
| Mean probability of infection$^{\dagger}$ (%) | 1.3 | 0.48 |
| Mean probability of infection$^{\ddagger}$, $\overline{P(R)}$ (%) | 3.2 | 9.8 |
| Proportion of population infected, $PPI$ (%) | 1.2 | 0.38 |
| Transmission ratio, $TR$ | 1 | 0.31 |

N, normal( $\mu, \sigma$ ); † , all spaces; ‡ , spaces where infected people present.

**Figure 1:**
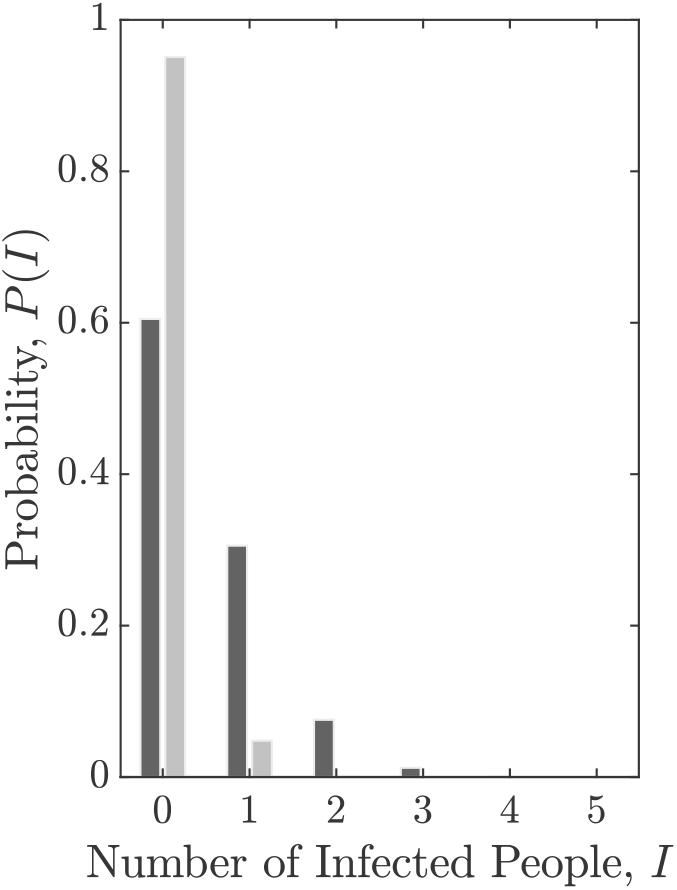
The probability of a number of infected people, *I*, present in the Big Office (dark) and Small Office (light), *P* (*I*), when *C* = 1%.

**Figure 2:**
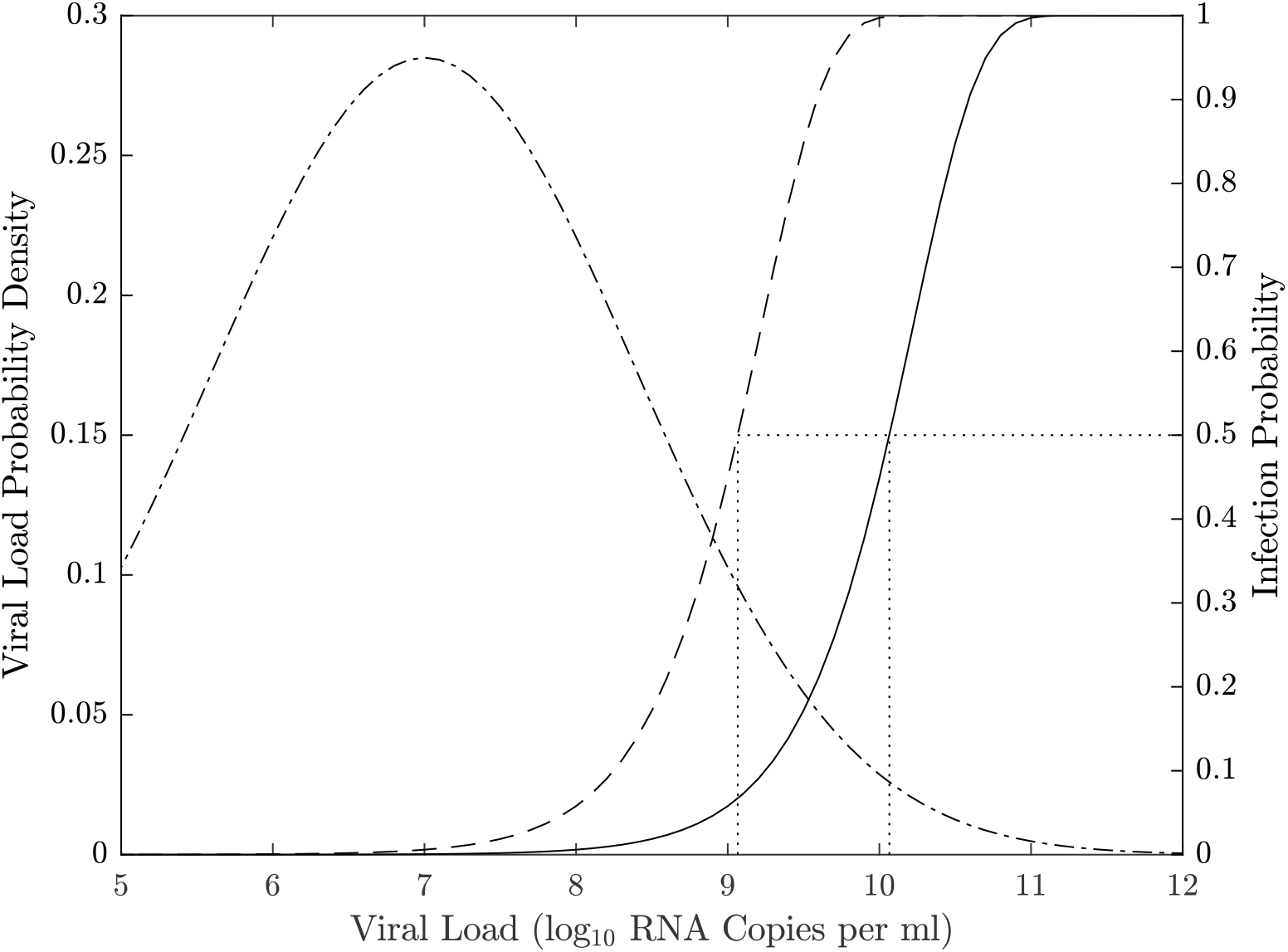
An indication of the relationship between the viral load, *L*, and the consequent probability of infection, *P* (*R*), in the Big Office (solid) and Small Office (dash) for a susceptible occupant, and the probability of a single infected person having a viral load, *P* (*L*), (dot-dash). Dotted vertical lines indicate the viral load required for *P* (*R*) = 50%.

### 2.2. Individual risk

A Relative Exposure Index (REI) is used to compare exposure risk for an individual person between two spaces following Jones *et al.* [15]. This approach has already been used to inform national policy on the role of ventilation in controlling SARS-CoV-2 transmission and to identify the appropriate application of air cleaning devices [24, 25].

The REI is the ratio of the dose, *D*, received by a susceptible occupant in each of two spaces using Equation 1 where the *reference* space is the denominator and the *comparator* space is the numerator. An advantage of using an REI is that uncertainty in the viral load of respiratory fluid ( RNA copies ml*^−^*^1^), which is used to determine the viral emission rate, *G* ( RNA copies m*^−^*^3^), and the unknown dose response both cancel allowing scenarios to be compared. When the REI is *>* 1 the comparator space is predicted to pose a greater risk to an individual susceptible occupant because they inhale a larger dose, although the absolute risk that this dose will lead to a probability of infection is not considered. Any space that wishes to have a REI of unity or less, must at least balance the parameters in Equation 1. A limitation of the REI is that it does not consider the probability of encountering an infected person with the same viral load in each scenario.

### 2.3. Population infection risk

The probability that a number of infected people, *I*, is present in a space, *P* (*I*), as a function of the number of occupants, *N*, is determined by considering the community infection rate, *C*, and standard number theory for combinations.

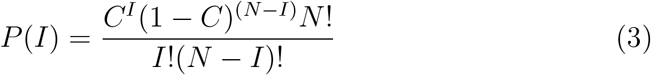

When a large population of people, *N_pop_*, is divided into a number of identical spaces, the total number of transmissions, *N_t_*, that occur is the sum of the number of transmissions that occur in each space.

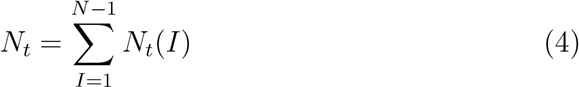

where *N_t_*(*I*) is the number of transmissions that occur in spaces that contain *I* infected people. For a large population, the number of people infected in each space is the product of the number of susceptible people exposed, *N_s_*, and the mean individual probability of infection in each space, 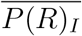.

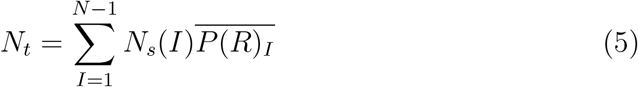

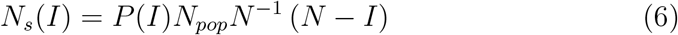

where *N_s_*(*I*) denotes the number of susceptible people exposed in spaces that contain *I* infected people, *P* (*I*) is the probability that a space contains *I* infected people, and *N_pop_ N^−^*^1^ denotes the total number of spaces that occur when a population *N_pop_* is divided into groups of *N* people. Here, the proportion of the population newly infected is given by

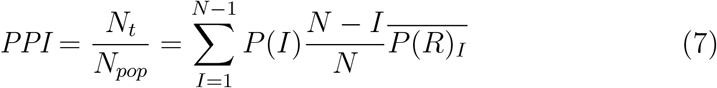

The exact solution for Equation 7 becomes increasingly difficult to evaluate as the space size increases. The calculation complexity is unlikely to be justified given the uncertainties in both the modelling assumptions and the available data. Therefore, simple approximations to the equation are desirable.

One approach is to express the number of transmission events using a single mean individual risk for all possible transmissions. Here, the *PPI* can be expressed as

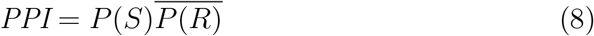

where *P* (*S*) is the proportion of the population who are both exposed and susceptible, and 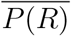 is the average individual infection risk that occurs in all spaces where there is a potential transmissions.

Transmission events can only occur when there are both one or more infected people present in a space (*I >* 0) and one or more susceptible people are present (*I < N*). It follows that the probability of a space containing a potential transmission event is given by

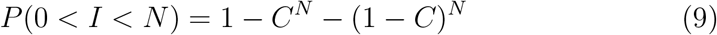

As the number of occupants tends to infinity, the probability that the space contains a potential transmission event approaches one, and is equal to zero for single occupancy spaces. This suggests that it may be better to partition a large space; see Section 1. Each space contains (*N − I*) susceptible people and the probability that an occupant is both susceptible and exposed is the difference between the number of susceptible people in the wider population, (1 *− C*) *N_pop_*, and the number of susceptible people who are not exposed, *P* (0) *N_pop_*. Therefore, *P* (*S*) is given by

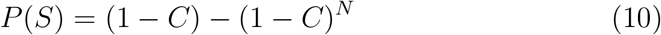

This equation shows that *P* (*S*) approaches the proportion of susceptible people in the wider population as *N → ∞*. *P* (*S*) can be minimised by reducing the community infection rate.

Evaluating the mean individual risk is non-trivial. Here an approximation is used, where

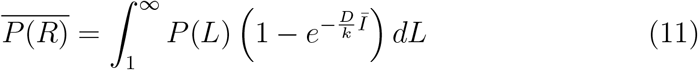

Here, *P* (*L*) is the probability of an infected person having a viral load *L*, and *Ī* denotes the mean number of infected people in a space that contains a potential transmission event, and is given by

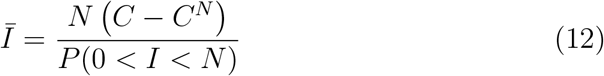

This allows the proportion of people infected in a scenario to be approximated by

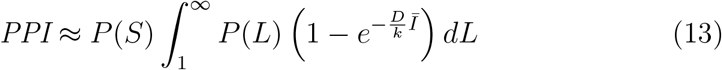

A transmission ratio, *TR*, gives an indication of the relative risk of infection between a *reference* and a *comparator* space where

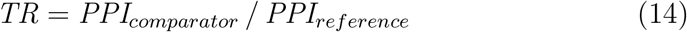

### 2.4. Scenarios

The probabilities given in Section 2.3 can be used to consider how the number of occupants may affect the relative exposure risk at population scale. First, we define a reference space against which others are compared. This space is an office, which is chosen because it is common and well regulated in most countries with consistent occupancy densities. The reference space has an occupancy density of 10 m^2^ per person, a floor to ceiling height of 3 m, and an outdoor airflow rate of 10 l s*^−^*^1^ per person. There are 50 occupants who are assumed to be continuously present for 8 hours breathing for 75% of the time and talking for 25%. Hereon it is known as the Big Office.

Then, we define a comparator space by subdividing the 50 person office into 10 identical spaces. Each space preserves the occupancy density, the *per capita* space volume, the outdoor airflow rate per person, and the air change rate. Hereon each comparator space is known as the Small Office.

All scenario inputs are given in Table 1.

### 2.5. Probabilistic estimates

A Monte Carlo (MC) model is used to corroborate the theory given in Section 2.3 and to investigate overdispersion in the model in Section 5.1. Pseudocode is given in Appendix B and MATLAB code is available under a creative commons license contained within the Supplementary Materials^1^

A population of 1 *×* 10^7^ people is divided into a number of identical spaces, which varies depending on the scenario; see Section 2.4 and Table 2. The population size is chosen so that the values of *PPI* and *TR*, rounded to two significant figures, do not change when the MC code is rerun. A binomial distribution can be used to model the number of successes in a number of independent trials, and so it is used to model both the number of infected people in each space and the number of susceptible people who are then infected when they inhale a dose of the virus. All inputs are given in Tables 1 and 2.

Uncertainty in other inputs are not explored because this has been done before [15] and to focus this work on an exploration of uncertainty in the viral load and the community infection rate.

## 3. Individual risk

The REI is the ratio of the dose predicted using Equation 1 for Big Office and Small Office; see Section 2.2. When the number of infected people and their respiratory activities, and the breathing rates of susceptible occupants, are identical in each space, the REI simplifies to a ratio of equivalent ventilation rates, *φ V*. The equivalent ventilation rate is used to determine the steady state concentration of viable virions. Table 1 shows that the removal rate *φ* is identical in both spaces and so the REI becomes a simple ratio of the number of occupants. This suggests that, in the presence of a single infected person, the relative risk is 10 times higher in the Small Office. This occurs because the Small Office contains ten times fewer people than the Big Office, and therefore the ventilation rate *per infector* is ten times smaller.

The equivalent ventilation rate per person, *φ V N^−^*^1^, is identical in both spaces and, if it is desirable to preserve the equivalent ventilation per person in two different spaces, the space volume per person must be preserved.

The removal rate, *φ*, includes the biological decay of the virus and the deposition of aerosols onto surfaces. Both of these removal mechanisms are space-volume dependent, and so their contribution to the removal of the virus is greater in spaces with a larger volume. Therefore, increasing the space volume per person also has the effect of reducing the REI. This has obvious physical limitations and a simpler approach is to reduce the number of people per unit of volume.

Equation 1 is used to calculate the dose of viable virions in each space and Table 1 shows that the magnitudes of the doses are small. There is great uncertainty in these values, attributable to modelling assumptions and in the inputs given in Table 1, but an increase by an order of magnitude still leads to a small dose. This fact is compounded by the value of unity for the viable fraction, which has the effect that all RNA copies inhaled are viable, which is unlikely. A viable fraction of unity was chosen because its true value is currently unknown, and this assumption simplifies the analysis. The value is clearly likely to be *«* 100% in reality, and so the actual doses would be substantially lower than those estimated here. This suggests that far-field transmission in buildings requires high viral emission rates, *G*, which are likely to be a rare event.

The probability of an infection occurring when a susceptible occupant is exposed to the dose reported in Table 1 is estimated using Equation 2 to be *P* (*R*) *<* 1% for both spaces and is approximately 10 times greater in the Small Office; see Table 2. Generally, this shows that the viral load has to be greater in the Big Office than in the Small Office to achieve the same *P* (*R*) when *C <* 1%. This is demonstrated by Figure 2, which describes the relationship between the viral load in respiratory fluid ( RNA copies ml*^−^*^1^) in each space attributable to any number of infected people and the consequent *P* (*R*) for a susceptible occupant, if the virus emission rate, *G*, is assumed to be linearly related to the viral load, *L*, of the infected person.

For any viral load the dose is calculated using Equation 1, and the probability that it leads to an infection is calculated using Equation 2. This creates a dose-response curve for both scenarios where factors that influence the REI and, therefore, the dose, determine the viral loads necessary to lead to a specific probability of infection. It also shows the relationship between the viral load and the probability that a single infected person has that viral load, *P* (*L*). The dotted vertical lines show the viral load required to give a 50% probability that the dose will lead to an infection for each scenario, *P* (*R*) = 50%. The area under the viral load probability density curve to the right of each vertical line is the probability that the viral load of the infected person leads to *P* (*R*) *≥* 50%. The probability is much smaller for the Big Office, which has the lower REI. This probability that an infected person has a viral load that leads to *P* (*R*) *≥* 50% is small, suggesting that the most likely outcome is *P* (*R*) *≤* 50%. There is great uncertainty in the magnitude of these values, particularly in *P* (*R*) and in the conversion of a viral load to a virus emission rate (see Section 2), but significant increases in them do not change the general outcomes of the analysis. More generally, increasing the number of occupants in a space while preserving the *per capita* volume has the effect of moving the *P* (*R*) curve to the right in Figure 2 and towards the tail of the *P* (*L*) curve, which reduces the likelihood that infected people in the space have a sufficient viral load.

The *P* (*L*) distribution curve could be flattened and shifted to the left of Figure 2 by reducing the viral load of the infected population. For example, vaccination is shown to clear the virus from the body quicker in infected vaccinated people, which at a population scale could flatten the distribution of *P* (*L*) [26]. However, different variants of the SARS-CoV-2 virus could increase the viral load, or the proportion of viable virions, or the infectivity of virions, and move the curve to the right of Figure 2 [27, 28]. Other respiratory viruses have different distributions of the viral load but the principles described here can be applied to them too.

## 4. Population risks

The analysis in Section 3 is underpinned by the assumption that there is a single infected person in each space. When the community infection rate (*C*) is known, Equation 3 can be used to estimate the probability that a specific number of infected people are present. Figure 1 shows that when *C* = 1%, in the Big Office *P* (*I* = 0) = 61%, *P* (*I* = 1) = 31%, and *P* (*I >* 1) = 9%. For the Small Office, *P* (*I* = 0) = 95%, *P* (*I* = 1) = 5%, and *P* (*I >* 1) is negligible. This shows that the Big Office is 8 times more likely to have an infected person present than the Small Office, although Table 1 shows that the relative risk is 10 times smaller in the Big Office than the Small Office when a single infected person is present. However, it is much more likely that both spaces do not have an infected person present, but when they are, the most likely number of infected people is 1. Equation 12 shows that the mean number of transmissions is *Ī* ≥ 1 for both scenarios when *C* = 1%.

Figure 2 shows the relationship between the probability of infection and the probability of a person having a particular viral load. The viral load that leads to an infection can be attributed to any number of infected people, but the probability of having more than 1 infected person in a space is generally small unless *N > C^−^*^1^; see Equation 9. When only 1 infected person is assumed to be present, Figure 2 also shows that the most probable viral loads are highly unlikely to lead to an infection in either the Small Office or the Big Office. Therefore, the infected person must have a significant viral load to infect susceptible occupants, which is an improbable event. The infection risk for susceptible occupants is lower in the Big Office than the Small Office when only 1 infected person is present.

Bigger spaces that preserve the *per capita* volume given in Table 1, and where *N »* 50, have a higher probability of susceptible people, *P* (*S*), and infected people, *P* (0 *< I < N*). The effect on the aerosol concentration and the dose depends on the space volume per infected person, *V I^−^*^1^, relative to that of the Reference Space, the Big Office. If *V I^−^*^1^ decreases, then the aerosol concentration, the dose, and the probability of infection, *P* (*R*), all increase. Accordingly, spaces with a high volume per occupant have a lower infection risk. Here, spaces with high ceilings or low occupancy densities are advantageous.

An increase in *C* also increases the probabilities of the presence of infected people, *P* (0 *< I < N*), and susceptible people, *P* (*S*), in any space. This increases the total viral load, the dose, *D*, and the probability of infection, *P* (*R*). Accordingly, maintaining a low community infection rate is important. It is worth noting that *C* may vary by region, or by a particular population demographic [29, 30]. Then, it is appropriate to use *C* for that demographic, rather than using a national value. It is possible to assess *C* by taking randomised samples from the population, such as the UK Coronavirus (COVID-19) Infection Survey [31], which includes all infected people at all stages of the disease. However, this survey includes symptomatic people who are likely to be isolating and so the actual *C* is likely to be lower.

The information in Figure 2 can be combined to determine the total proportion of people newly infected, *PPI*, in a space for all viral loads as a function of the probability that an individual infected person has a particular viral load, *P* (*L*), the probability of the risk of infection, *P* (*R*), the probability of the presence of susceptible people *P* (*S*), and the average number of infected people, *Ī*; see Equations 7 and 8.

Figure 3 shows the relationship between the PPI and the viral load where the area under each curve is the proportion of the entire population infected when *C* = 1% and assuming that two equal populations are each distributed evenly across a number of spaces; the first across a number of Big Office spaces and the second distributed across a larger number of Big Office spaces. The area under the curve and thus the values for the population PPI are confirmed using the MC analysis described in Section 2.5 and given in Table 2. Table 2 indicates that the probability of far-field infection is *PPI* = 0.38% in the Small Office and *PPI* = 1.2% in the Big Office. The *TR* is calculated using Equation 14 and is 0.31. Therefore, the infection risk is 3 to 4 times higher in the Big Office.

**Figure 3:**
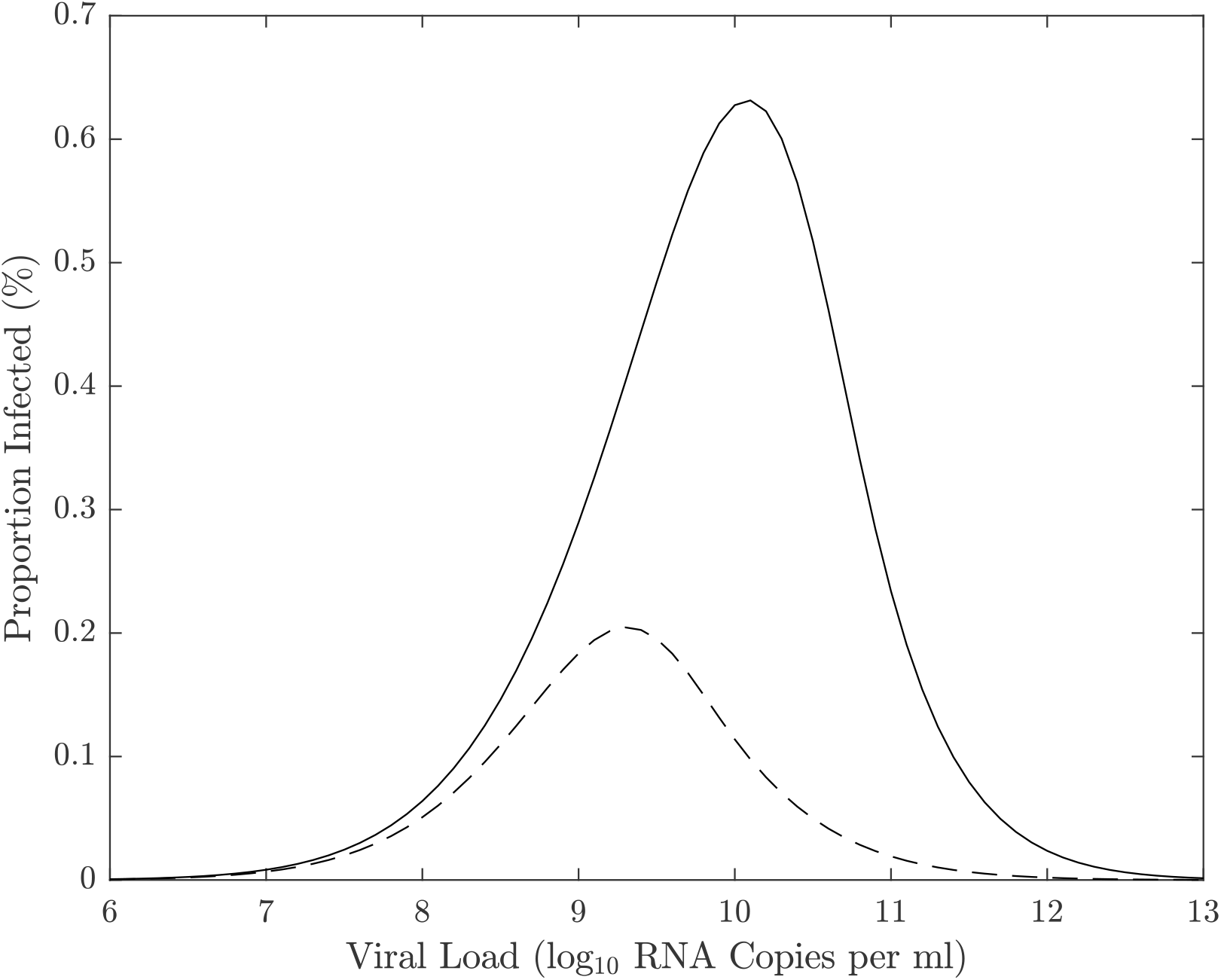
An indication of the relationship between the proportion of a population infected for a particular viral load when the community infection rate is *C* = 1%. The area under the curve represents the total proportion of people infected for the Small Office (dash) and the Big Office (solid).

The absolute values of PPI are likely to be much smaller than those calculated here because of the conservative assumptions used to estimate the viral emission from the viral load (see Section 2.1), so the *PPI* may well be *«* 1% in both spaces using less conservative assumptions; see the Supplementary Materials^1^. This indicates that although there are benefits of subdividing for a population, their magnitude needs to be considered against other factors, such as the overall work environment, labour and material costs, and inadvertent changes to the ventilation system and strategy.

The uncertainties in all of the values given here are significant and so it is not possible to be confident in the magnitude of the *PPI* or the *TR*, but testing the model with a range of assumptions enables an assessment of general trends; for example, how increasing occupancy and preserving *per capita* space volume and ventilation rates impact the risk of infection and how different mitigation measures, such as increasing the ventilation rate, affect the relative *PPI*. These are discussed in Section 5.

## 5. Discussion

### 5.1. Overdispersion

The MC approach described in Section 2.5 was used to corroborate the mathematics given in Section 2.3. The predictions given in Table 2 can be produced using either method, giving confidence in the concept and the model.

The MC approach is used to interrogate each space and estimate the number of susceptible people infected in the Big Office, when an infected person is present. The proportion of the susceptible population infected in each space is given in Figure 4. It predicts that there were no transmissions in 90% of the spaces. However, when a transmission does occur, the most common outcome is a single transmission event. This indicates that the dose inhaled by all susceptible people is usually small enough not to lead to an infection. This is confirmed by Figure 5, which gives the cumulative distribution of dose for both scenarios. It shows that susceptible occupants receive no dose in Big Office spaces 61% of the time and 95% of the time in Small Office spaces.

**Figure 4:**
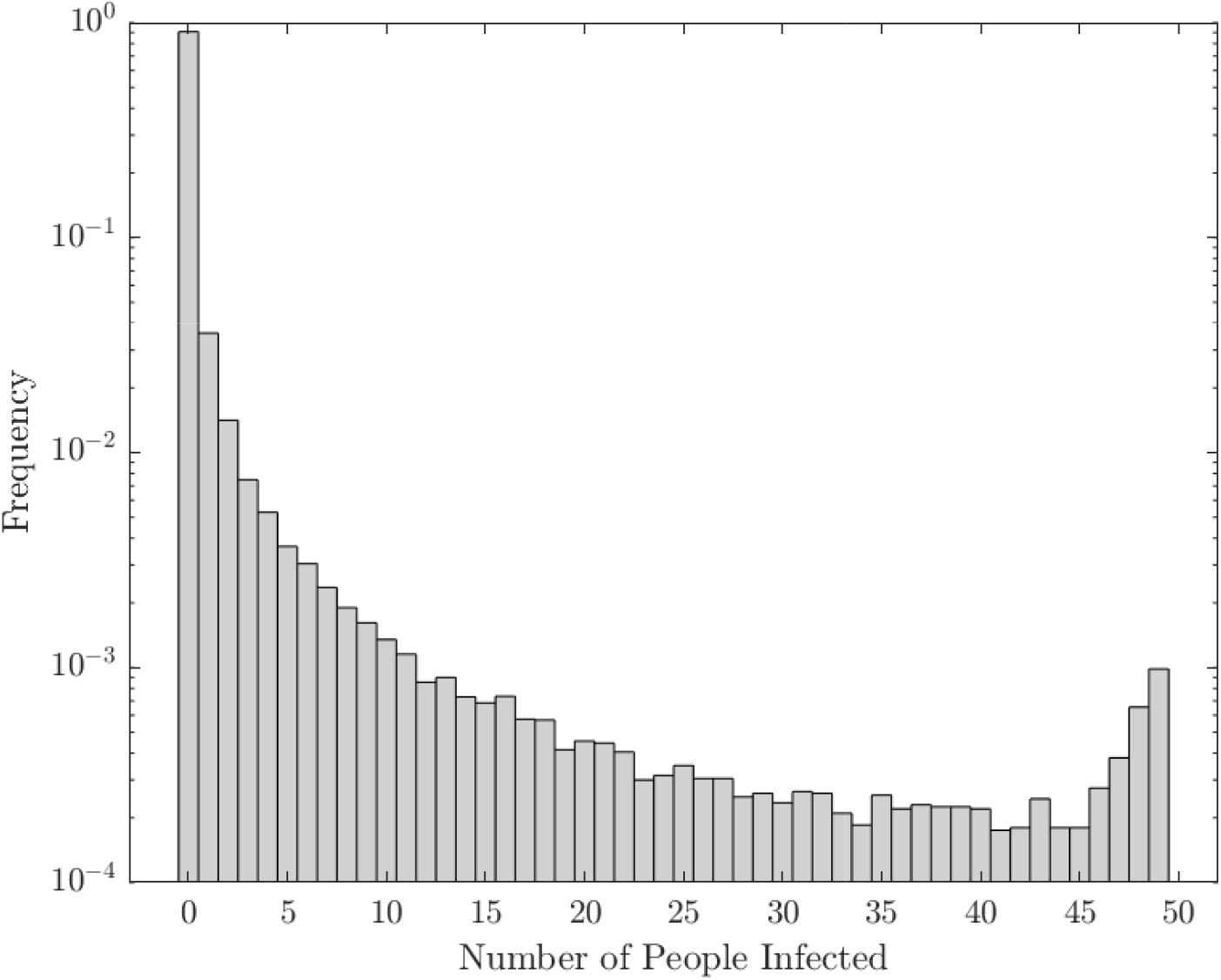
The number of susceptible people infected in each Big Office space estimated using a Monte Carlo approach.

**Figure 5:**
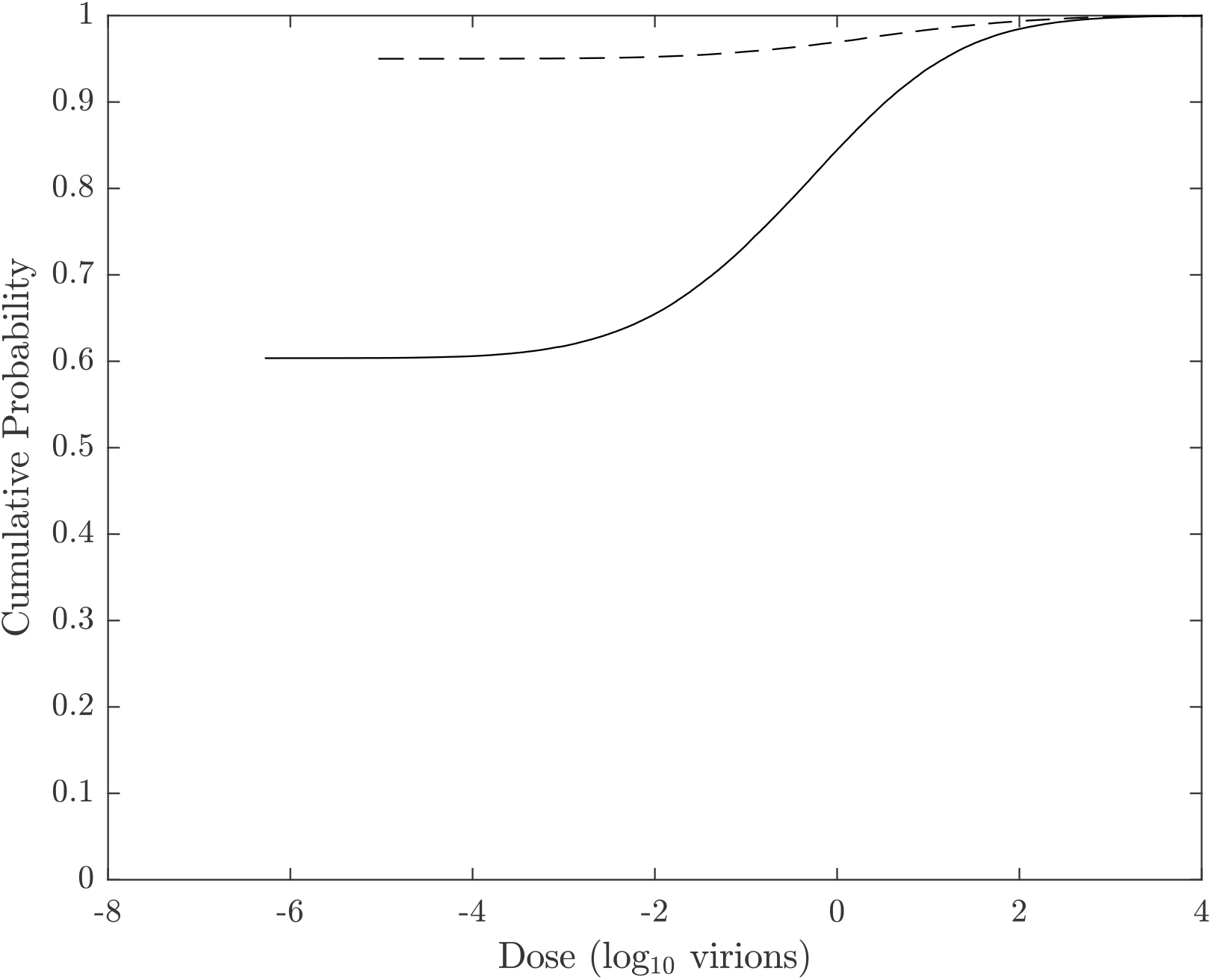
The cumulative probability of the dose in the Big Office (solid) and the Small Office (dashed) when *C* = 1%.

More than 40 susceptible people are infected in the Big Office only 0.3% of the time; see Figure 4. This suggests that so called *super-spreader* events that occur by far-field airborne transmission alone, are likely to be rare. This distribution reflects the overdispersion of transmission recorded for SARS-CoV-2 and, although this work only considers one transmission route, similar relationships between the viral load and the number of transmission events may also be true for other transmission routes [11, 32, 33, 34, 35, 36, 37, 38]. Applying the MC approach to the Small Office shows that the overdispersion is less pronounced because there are fewer susceptible people and fewer infected people in each space. This limits the number of susceptible people who can be infected when the viral load is high. Here, 0.2% of all spaces, and 22% of spaces with at least one transmission, had 4 infections of susceptible people.

There are very few epidemiological examples of high secondary COVID-19 transmission events where *>* 80% of occupants in a space are infected and this suggests that our assumptions over-estimate the viral emission rate. One reason is the assumption that all genome copies are viable virions, which is very unlikely.

Figure 4 shows that the frequency of the number of susceptible people infected is highest at zero and decreases as the number of susceptible people infected increases. However, the frequency later increases as the number of susceptible people infected approaches the number of occupants. This reflects the shape of the probability of infection curve in Figure 2 where a point is reached when the viral load leads to the infection of all susceptible people, and a higher viral load cannot infect more people. The phenomena is a function of occupancy and is less likely to occur as the number of occupants increases because the viral load required to infect all susceptible people increases, assuming that the *per capita* space volume and ventilation rate are constant.

### 5.2. Ventilation and space volume

The quotient of the proportion of people infected in the two scenarios gives a Transmission Ratio, *TR*, see Equation 14. Increasing the *per capita* ventilation rate, *ψ V N^−^*^1^, or space volume, *V N^−^*^1^, in the Big Office reduces the inverse of the *TR*. This has the effect of increasing the total removal rate, *φ*, and reducing the dose and the probability of infection; see Equation 1 and Figure 6. However, there is a law of diminishing returns in reducing the *PPI* by increasing the ventilation rate because the dose is inversely proportional to *φ*. Therefore, it is more important to increase the ventilation rate in a poorly ventilated space than in a well ventilated space because the change in the *PPI* is greater.

**Figure 6:**
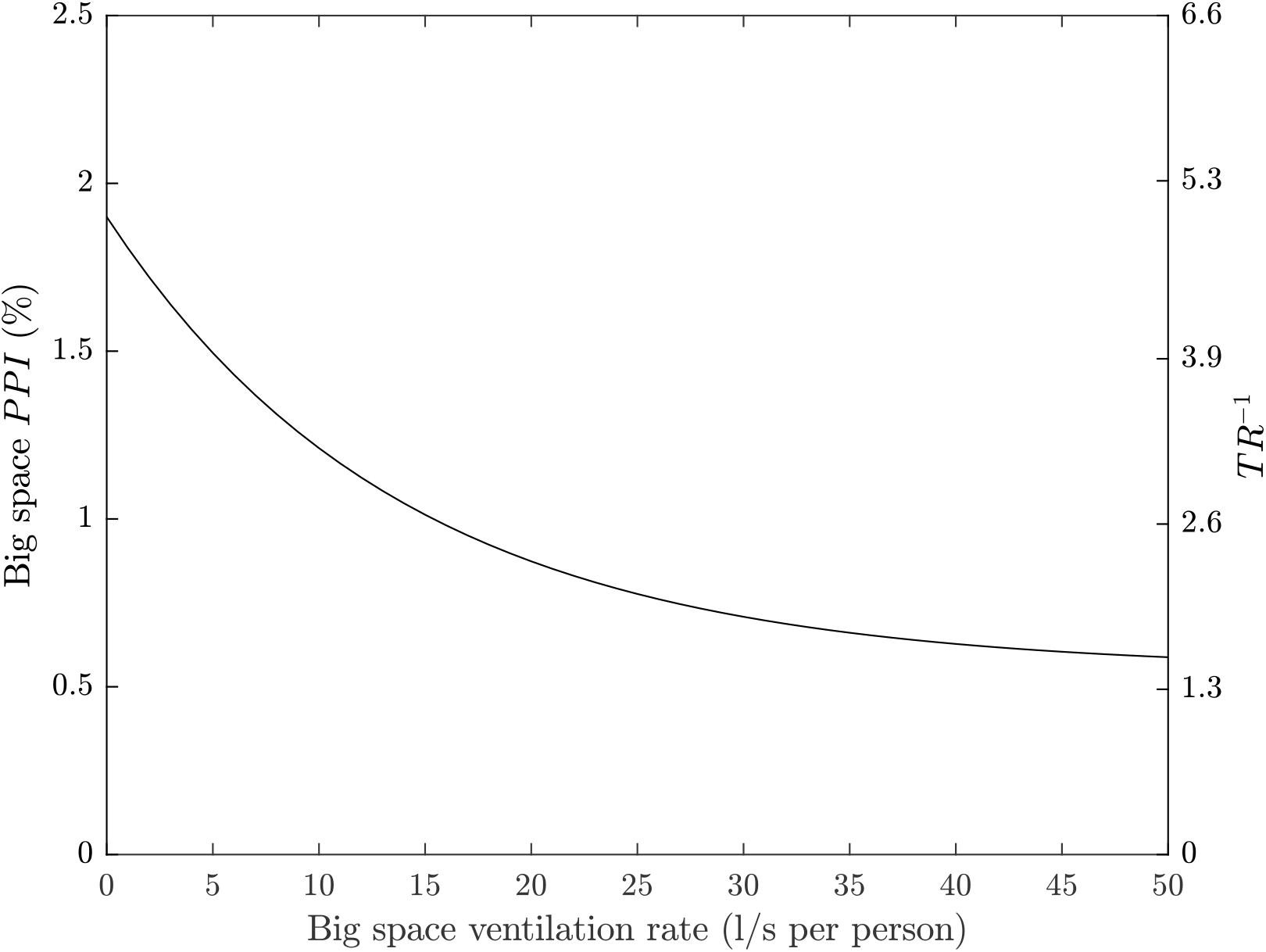
The effect of increasing the *per capita* ventilation rate, *ψ V N^−^*^1^, in the Big Office on the *PPI* and the *TR* when the *per capita* ventilation rate in the Small Office is a constant 10 l s*^−^*^1^ per person. All values are illustrative.

A similar effect is seen when increasing the *per capita* space volume in the Big Office while maintaining a constant *per capita* ventilation in both spaces. This is because the dose is inversely proportional to volume. Furthermore, the product of the space volume and the total removal rate, *φ V*, is proportional to the concentration of the virus in the air and, therefore, the infectious dose. The *per capita* ventilation rate is constant in both spaces and so the air change rate in the Big Office decreases as its volume increases. However, this reduction is offset by the surface deposition and biological decay rates, which remain constant and have a greater effect on the value of the equivalent ventilation rate, *ψ V*, as the space volume increases; see Section 2.1.

Equation 1 assumes a steady-state concentration of the virus has been reached based on the assumption that the exposure time, *T*, is significant. However, the time taken to reach the steady-state concentration in large spaces may be significant and affects the dose over shorter exposure periods. This is an example of the *reservoir effect*, the ability of indoor air to act as a fresh-air reservoir and absorb the impact of contaminant emissions. The greater the space volume, the greater the effect. These factors highlight the benefits of increasing the *per capita* space volume.

### 5.3. Occupancy

Figure 7 shows the effect of increasing the number of occupants in the Big Office while maintaining both the *per capita* space volume, *V N^−^*^1^, and ventilation rate, *ψ V N^−^*^1^. As the number of occupants increases, the *PPI* increases at an ever diminishing rate because the magnitude of the equivalent ventilation rate, *φ V*, increases at a greater rate than the probability of the mean number of infected people, *Ī*.

**Figure 7:**
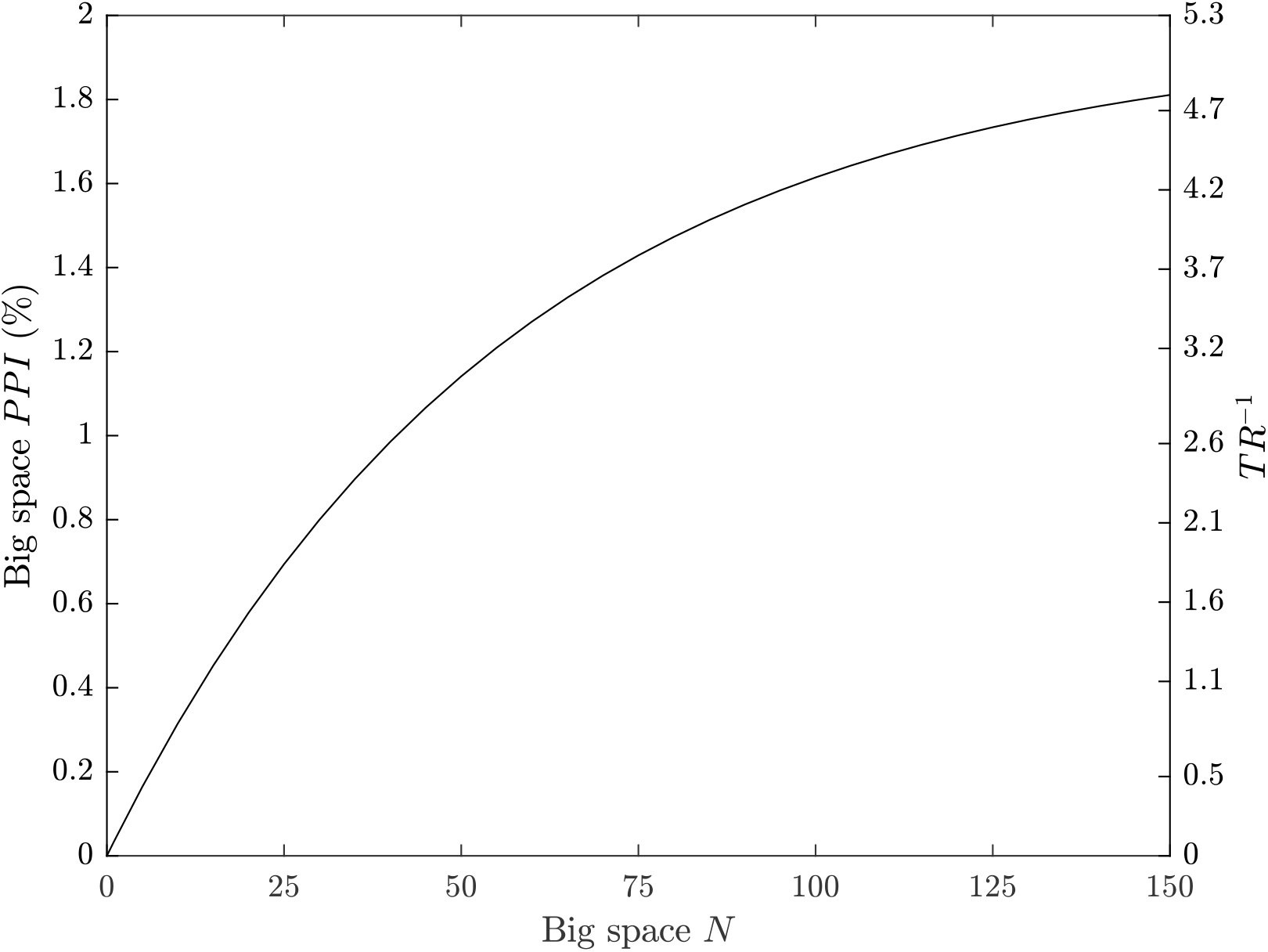
The effect of increasing the occupancy, *N*, in the Big Office, where the space volume per person and ventilation rate per person is fixed at 30 m^3^ and 10 l s*^−^*^1^ respectively, on the *PPI* (green) and *TR* (black). All values are illustrative.

However, if the volume and ventilation rate remain constant as the occupancy increases, Figure 8 shows that the *PPI* and the inverse of the *TR* increase linearly with occupancy. Here, the total removal rate, *φ*, remains constant but the *per capita* space volume and ventilation rate reduce. Therefore, the Big Office could have 14 occupants and have the same *PPI* as the Small Office occupied by 5 people. Extrapolating to two identical populations of 140 people split into 28 Small Offices with 5 people in each, and 10 Big Offices with 14 people in each, the same *PPI* can be achieved.

**Figure 8:**
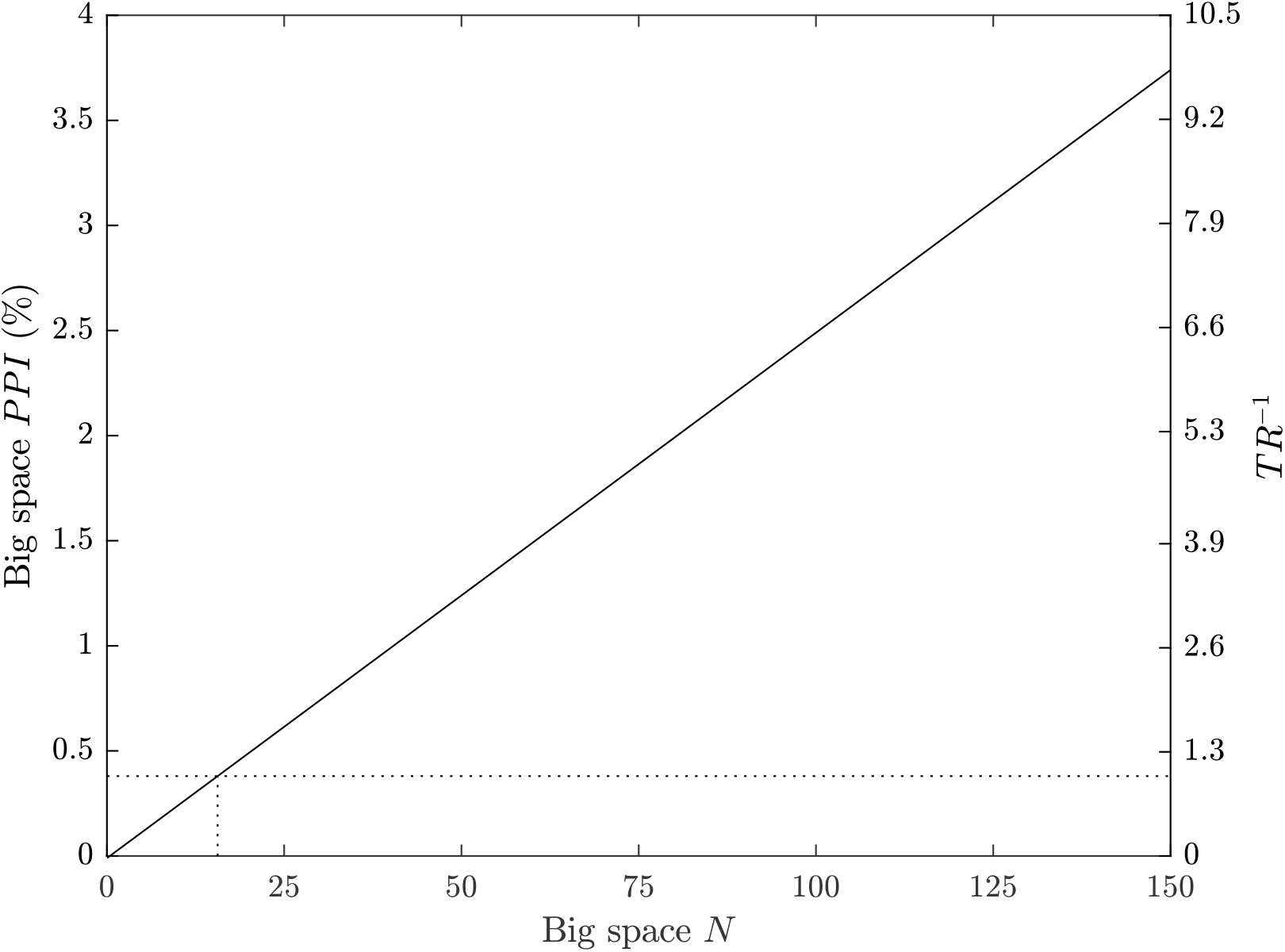
The effect of increasing the occupancy, *N*, in the Big Office where the space volume and ventilation flow rate are fixed for a designed occupancy of 50 people (1500 m^3^ and 500 l s*^−^*^1^, respectively), on the *PPI* and *TR*. All values are illustrative.

This suggests that reducing the number of occupants in a space is the most effective means of reducing the inverse of *TR* towards unity. To achieve the same goal by increasing the ventilation rate or the *per capita* space volume would require unfeasibly large increases in both.

### 5.4. Community infection rate

Figure 9 shows that the community infection rate, *C*, has a significant effect on the *PPI* and the *TR*. This is because it affects both the probability of an infectious level of viral load, *P* (*L*), and the probability of having susceptible people in a space, *P* (*S*); see Equation 10. When *C >* 1%, the probability of transmission increases dramatically, suggesting that it strongly influences the spread of the virus indoors. Figure 9 also shows that *C* only affects the *TR* when the number of occupants, *N*, is less than the reciprocal of the community infection rate in both spaces, *N <* 1*/C*. Thereafter, the *TR* is constant irrespective of the community infection rate; see the Supplementary Materials^1^.

**Figure 9:**
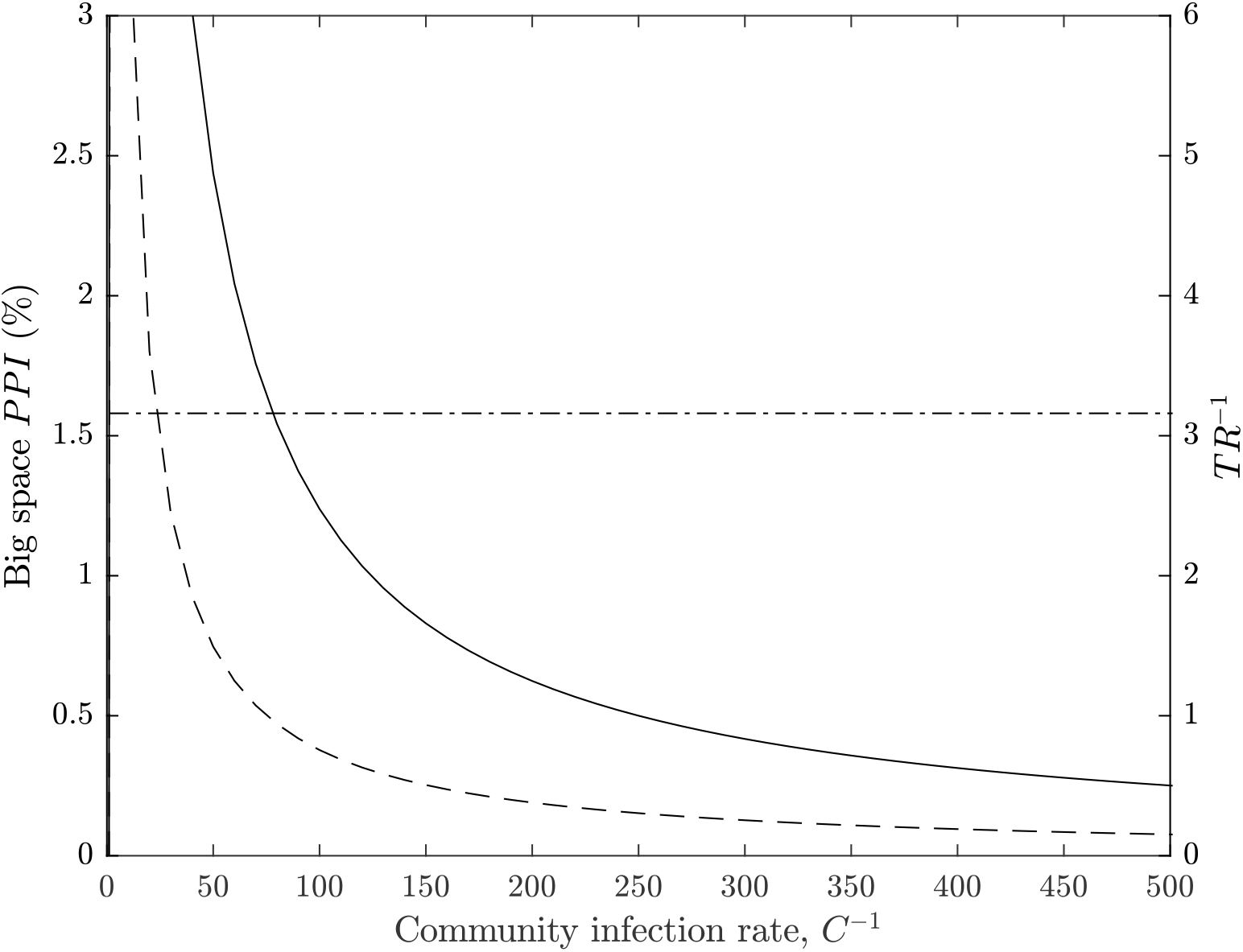
The effect of decreasing the community infection rate, *C*, on the *PPI* in the Big Office (solid) and the Small Office (dash) and on the *TR* (dot-dash). All values are illustrative.

### 5.5. Limitations

Some limitations and uncertainties in this work have already been addressed, particularly those concerning the viral load and the dose-response relationship. However, there are a number of other aspects that increase uncertainty in it. Firstly, the models assume homogenous instantly mixed indoor air to simplify the estimate of a dose. This assumption is unlikely to be true in some spaces, especially in large spaces where the concentrations of virions in the air is likely be a function of the distance from the infected person, although it is unclear at which space volume this assumption becomes less useful[39].

The approach described in Section 2 only considers the far-field transmission of virus, and not near-field transmission, which is likely to be the dominant route of transmission. The concentration of the virus in aerosols and droplets per unit volume of air is several orders of magnitude greater closer to the infected person at distances of *<* 2 m [3, 9]. However, it is likely that the method of calculating the probability of viral load of infected people, *P* (*L*), is also important for the dose received by near-field transmission and should be explored further in the future.

The distribution of viral load of an infected person around the median will affect the probability of transmission. We apply a normal distribution of log_1_0 values, see Section 2, but another, such as the Weibull distribution, will affect the transmission probabilities differently.

The model also assumes a näıve population of susceptible people, and it is unclear whether a higher infectious dose is required for susceptible people who have a greater immune response obtained from vaccination or a previous infection. It also assumes everyone is equally susceptible, which is unlikley. This paper does not consider the effect of the magnitude of the dose on subsequent disease severity. However, a recent review suggests that it is highly unlikely there is a link between dose and disease severity [40].

There is uncertainty in the dose-response relationship and the proportion of people infected. In the absence of knowledge, we have assumed that the dose-response curve for SARS-CoV-1 also applies to SARS-CoV-2; see Section 2.1. The SARS-CoV-1 dose-response curve was generated from four groups of inoculated transgenic mice [23] that were genetically modified to express the human protein receptor of the SARS-CoV-1 virus. In three of the groups all mice were infected and in the fourth one-third were infected. The dose-response curve was fitted to data from these four groups and, although it is limited, it is sufficient to assume that the curve follows the exponential distribution rather than the Beta-Poisson distribution. A further limitation is that the response of humans to a dose of SARS-CoV-1 may vary significantly from that of transgenic mice. For a further discussion, see the Supplemental Material^1^. There is also uncertainty in the measurement of the viral load used to challenge the study, and whether or not dose curves are valid for predicting low probabilities of infection at very low virus titres. Other studies have used alternative dose-response curves for other coronaviruses, all of which have similar uncertainties [21, 16], but this framework provides a means to test other dose-response relationships by adjusting *k* in Equation 1.

The viral load of an infected person is the number of RNA copies ml*^−^*^1^ of respiratory fluid, whereas the viral emission is the amount of RNA copies per unit volume of exhaled breath; see Section 2.1. It has been established that the viral load of an infected person increases in time from the moment of infection and is highest just before, or at, the onset of COVID-19 symptoms. As COVID-19 progresses the viral load reduces, normally within the first week after the onset of symptoms [41, 42]. The viral load also varies between people at any stage of the infection, which increases uncertainty in it [43, 44, 45, 19, 46, 18, 30, 37, 47].

The viral load can be inferred from the *cycle threshold* values of real time reverse transcription quantitative polymerase chain reaction (RT qPCR) nasopharyngeal (NP) swabs. This method assumes a direct correlation between the viral load of a swab and the viral load of respiratory fluid [48, 12]. RT qPCR is a semi-quantitative method because it requires a number of amplification cycles to provide a positive signal of the SARS-CoV-2 genome, which is proportional to the initial amount of viral genome in the original sample. The cycle threshold is the number of polymerase chain reaction cycles that are required before the chemical luminescence is read by the equipment. The lower the starting amount of viral genome, the greater the number of amplification cycles required. A calibrated standard curve is then used to estimate the starting amount of viral genomic material. However, the standard curve varies between test assays (investigative procedures) and different RT qPCR thermal cyclers, the laboratory apparatus used to amplify segments of RNA. This method also assumes a complete doubling of genetic material after each cycle. The exponential relationship means that errors in the calculation of the initial quantity of genomic material are orders of magnitude higher for low cycle counts than for high cycle counts. Additionally, if genomic data is taken from NP swabs, the estimated concentration of genomic material per unit volume is often related to the amount of genomic material in the buffer solution^2^ in which NP swabs are eluted and used in the assay, and not necessarily to the amount in a patient’s respiratory fluid. The amount of genomic material added to the buffer solution is dependent on both a patient’s viral load and the quality of the collection of the NP sample, which is highly variable. Therefore, it is not possible to determine absolute values of the viral load in a patient’s respiratory fluid using this method. However, data collected in this way is indicative of a range of variability, much of which is likely to be proportional to the viral load of the person at the time the sample was collected. Some recent data suggests that the viral load of NP swabs may not reflect the amount of infectious material present [19]. However, it is important to note that there are wide variations in the measured genomic material in NP swabs and that the viral load in respiratory fluid is likely to vary by several orders of magnitude.

There is clearly uncertainty in the viral load of respiratory fluid. There is also uncertainty in the viral concentration in respiratory aerosols and droplets and the distribution is currently unclear. Some studies suggest that the number of virions in small aerosols with a diameter of *<* 1 *µ*m is higher than would be expected given the viral concentration in the respiratory fluid [49, 50] and that for SARS-CoV-2 there may be more genomic material in the smallest aerosols [51].

There is high variability between people in the total volume of aerosols generated per unit volume of exhaled breath, and it is dependent upon the respiratory activity, such as talking and singing, and the respiratory capacity [52, 53, 54, 55]. Coleman *et al.* [51] show that SARS-CoV-2 genomic material is detectable in expirated aerosols from *some* COVID-19 patients, but not all of them because 41% exhaled no detectable genomic material. Singing and talking generally produce more genomic material than breathing, but there is large variability between patients. This suggests that respiratory activities that have previously been shown to increase aerosol mass also increase the amount of viral genomic material emitted. However, the viral concentration in aerosols cannot be determined because the study did not measure the mass of aerosols generated. Coleman *et al.* also show that the variability in the amount of genomic material measured in expirated aerosols is consistent with the variability of viral loads determined using swabs and saliva [51].

Similarly, Adenaiye *et al.* [56] detected genomic material in aerosols from patients infected with SARS-CoV-2 who provided a sample of exhaled air when talking or singing. Genomic material was more frequently detected in exhaled aerosols when the viral load of saliva or mid-turbinate swabs was high; *>* 10^8^ and *>* 10^6^ RNA copies for mid-turbinate swabs and saliva samples, respectively. Furthermore, they were able to culture viable virus from *<* 2% of fine aerosol samples. It should be noted that one positive sample was from a culture obtained from a fine aerosol sample that had an amount of genomic material that was less than the detection limit of the qRT PCR method, so it could be an artefact. Nevertheless, this provides some evidence to support the epidemiological evidence that viable virus can exist in exhaled aerosols.

Miller *et al.* suggests that around 1 : 1000 genome copies are likely to be infectious virion [57, 12]. Adenaiye *et al.* use mid-turbinate swabs to estimate that there are around 1 : 10^4^ viable virus per measured genome copies[56]. We make the assumption that all genome copies are viable virion, which either over-estimates their infectiousness when using the Coleman *et al.* data, or is similar to the assumption of Miller *et al.* if the viable virion emission rate (calculated from air in a hospital) is in the order of 1000 virions per hour; see Appendix A.

## 6. Conclusions

The number of occupants in a space can influence the risk of far-field airborne transmission that occurs at distances of *>* 2 m because the likelihood of having infectious and susceptible people are both associated with the number of occupants. Therefore, mass-balance and dose-response models are applied to determine if it is advantageous to sub-divide a large reference space into a number of identical smaller comparator spaces to reduce the transmission risk for an individual person and for a population of people.

The reference space is an office with a volume of 1500 m^3^ occupied by 50 people over an 8 hour period, and has a ventilation rate of 10 l s*^−^*^1^ per person. The comparator space is occupied by 5 people and preserves the occupancy period and the *per capita* volume and ventilation rate. The dose received by an individual susceptible person in the comparator Small Office, when a single infected person is present, is compared to that in the reference Big Office for the same circumstances to give a relative exposure index (REI) with a value of 10 in the Small Office. This REI is a measure of the risk of a space relative to the geometry, occupant activities, and exposure times of the reference scenario and so it is not a measure of the probability of infection. Accordingly, when a single infected person is assumed to be present, a space with more occupants is less of a risk for susceptible people because the equivalent ventilation rate per infected person is higher.

The assumption that only one infected person is present is clearly problematic because, for a community infection rate of 1%, the most likely number of infected people in a 50 person space is zero. A transmission event can only occur when there are both one or more infected people present in a space and one or more susceptible people are present. The probability of a transmission event occurring increases with the number of occupants and the community infection rate; for example, the Big Office is over 12 times more likely to have infected people present than the Small Office. However, the geometry and ventilation rate in a larger space are non-linearly related to the number of infected and susceptible people and so their relationship with the probability of a transmission event occurring is also non-linear. These effects are evaluated by considering a large population of people. But, this introduces uncertainty in factors that vary across the population, such as the viral load of an infected person, defined as the number of RNA copies ml*^−^*^1^ of respiratory fluid. The viral load varies over time and between people at any stage of the infection.

By applying a distribution of viral loads across a population of infected people, secondary transmissions (new infections) are found to be likely to occur only when the viral load is high, which agrees with Schijven *et al.*[38], although the probabilities of this occurring in the Big Office and the Small Office are low. This makes it hard to distinguish the route of transmission epidemiologically. Generally, the viral load must be greater in the Big Office than in the Small Office to achieve the same proportion of the population infected when the community infection rate is *≤* 1%. The viable fraction is unknown but a value of unity was chosen for computational ease, yet the estimated doses and infection probabilities are small. Therefore, it is likely that far-field transmission is a rare event that requires a high emission rate and that there is a set of Goldilocks conditions that are *just right* where ventilation is an effective mitigation method against transmission. These conditions depend on the viral load, because when it is low or high, ventilation has little effect on the risk of transmission.

There are circumstances where the magnitude of the total viral load of the infected people is too high to affect the probability of secondary transmissions by increasing ventilation and space volume. Conversely, when the total viral load is very small, the dose is so small that it is highly unlikely to lead to an infection in any space irrespective of its geometry or the number of susceptible people present. There is a law of depreciating returns for the dose and, therefore, the probability of infection, and the ventilation rate because they are inversely related. Accordingly, it is better to focus on increasing effective ventilation rates in under-ventilated spaces rather than increasing ventilation rates above those prescribed by standards, or increasing effective ventilation rates using air cleaners, in already well-ventilated spaces.

There are significant uncertainties in the modelling assumptions and the data used in the analysis and it is not possible to have confidence in the calculated magnitudes of doses or the proportions of people infected. However, the general trends and relationships described herein are less uncertain and may also apply to airborne pathogens other than SARS-CoV-2 at the population scale. Accordingly, it is possible to say that there are benefits of subdividing a population, but their magnitudes need to be considered against other factors, such as the overall working environment, labour and material costs, and inadvertent changes to the ventilation system and strategy. However, it is likely that the benefits do not outweigh the costs in existing buildings when a less conservative viable fraction or a lower community infection rate is used because it decreases the magnitude of the benefits significantly. It is likely to be more cost-effective to consider the advantages of partition when designing new resilient buildings because the consequences can be considered from the beginning.

There are other factors that will reduce the risk of transmission in existing buildings. Local and national stakeholders can seek to maintain low community infection rates, detect infected people with high viral loads using rapid antigen tests and support to isolate them (see the Supplementary Materials^1^), reduce the variance and magnitude of the viral load in a population by encouraging vaccination [30]. Changes can be made to the use of existing buildings and their services, such as reducing the occupancy density of a space below the level it was designed for while preserving the magnitude of the ventilation rate, reducing exposure times, and ensuring compliance with ventilation standards.

## Supporting information

Matlab code

## Data Availability

All data produced in the present work are contained in the manuscript and Supplemental Materials

## Acknowledgements

The authors acknowledge the Engineering and Physical Sciences Research Council (EP/W002779/1) who financially supported this work. They are also grateful to Constanza Molina for her comments on this paper.

## Appendix A. Estimating viral emission from viral load

We assume that the RNA copies ml*^−^*^1^ concentration is constant in aerosols and in NP swabs and then we use the assumptions of Jones *et al.* [15] to convert a NP viral load into a virus emission rate. This method follows Jones *et al.* and is derived from the work of Morawskwa *et al.* who determine volume distribution aerosols for different respiratory activities, and is similar to that used by Lelieveld *et al.* [15, 17, 55]. Table A.3 shows the estimated virus emission rate for different respiratory activities when the viral load is 10^7^ RNA copies ml*^−^*^1^. For comparison, median measured values of virus emission in aerosols from Coleman *et al.* are given. These values were measured by collecting RNA copies from COVID-19 patients, where the median cycle threshold, required to process diagnostic samples, was 16. [51].

**Table A.3:** Estimated emission rates from an infected person with a viral load of 10^7^ RNA copies ml*^−^*^1^ compared to measured emission rates from patients with a median cycle threshold of 16 [51]

|  | Estimated | Measured median |
| --- | --- | --- |
| | RNA copies $\text{h}^{-1}$ | RNA copies $\text{h}^{-1}$ |
| Breathing | 203 | 127 |
| Voiced counting (talking) | 967 | 1912 |
| Vocalisation (singing) | 6198 | 2856 |
| Breathing:talking 25:75 | 394 | 573* |

\*calculated using measured values for breathing and talking.

Additionally, unpublished work by Adenaiye *et al.* measured viral genome in patients infected by the SARS-CoV-2 alpha variant, who were breathing and talking, in coarse (*>* 5 *µm*) and fine (*≤* 5 *µm*) aerosols with a total geometric mean of 1440 RNA copies h*^−^*^1^ and a maximum of 3*×*10^5^ RNA copies h*^−^*^1^ [56]. These are greater than the estimated values given in Table A.3, but the viral load, measured by genome copies from mid-turbinate swabs, was generally orders of magnitude higher than 10^7^ RNA copies ml*^−^*^1^.

In Section 4, the inhaled dose is calculated for all possible viral loads. Here, it should be noted that the calculated RNA copies emission rate is assumed to be linearly related to the viral load of respiratory fluids, so that a viral load of 10^8^ RNA copies ml*^−^*^1^ has a ten-fold greater emission rate. For comparison, a virus emission rate of 394 RNA copies h*^−^*^1^ (assumed for a viral load of 10^7^ RNA copies ml*^−^*^1^) leads to individual doses of around 2.2 RNA copies and 0.2 RNA copies for the Small Office and Big Office scenarios, respectively.

The calculated emission rate of viral genome for a viral load of 10^7^ RNA copies ml*^−^*^1^ is a reasonable fit to the Coleman *et al.* and Adenaiye *et al.* data. For further details see the Supplementary Materials^1^.

## Appendix B. Pseudocode

~~~
SET population size
SET scenario space volumes
SET scenario people per space
FOR each scenario
    COMPUTE number of spaces
    FOR each space
        SAMPLE infected people from binomial distribution
        IF infected people is number of occupants THEN
           SET infected people to zero
        END IF
        COMPUTE susceptible & exposed people
        IF infected people is zero THEN
           SET susceptible & exposed people to zero
        END IF
        SAMPLE log10 viral load from normal distribution
        COMPUTE emission rate using viral load
        COMPUTE dose using emission rate
        COMPUTE probability of infection per susceptible person
        SAMPLE infected susceptible people from binomial distribution
    END FOR
    COMPUTE number of transmission events
    COMPUTE probability of infected people present
    COMPUTE individual probability being susceptible & exposed
    COMPUTE mean number of infected people
    COMPUTE mean emission rate
    COMPUTE mean dose
    COMPUTE mean probability of infection
    COMPUTE proportion of population infected
END FOR
COMPUTE transmission ratio
~~~

## Nomenclature

Ī: mean number of infected people in a space that contains a potential transmission event
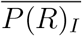: mean individual probability of infection occurring in each space
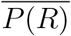: mean individual infection risk that occurs in all spaces with a potential transmission
*Φ*: total removal rate ( s*^−^*^1^)
*C*: community infection rate
*D*: dose (viable virions)
*G*: emission rate of RNA copies ( RNA copies s*^−^*^1^)
*I*: number of infected people
*K*: fraction of aerosol particles absorbed by respiratory tract
*K*: reciprocal of the probability that a single pathogen initiates an infection
*L*: viral load ( RNA copies ml*^−^*^1^ of respiratory fluid)
*N*: number of occupants
*N_s_*: number of susceptible people exposed
*N_s_*(*I*): number of susceptible people exposed in spaces that contain *I* infected people
*N_t_*: number of transmissions for an entire population
*N_t_*(*I*): number of transmissions that occur in spaces that contain *I* infected people
*N_pop_*: population size
*P* (0 *< I < N*): probability of a space containing a potential transmission
*P* (*I*): probability of *I* infected people present
*P* (*L*): probability of a viral load
*P* (*R*): individual infection probability for a given dose
*P* (*S*): probability of a person being both susceptible and exposed to the virus
*PPI*: proportion of a population infected
*q_resp_*: respiratory rate ( m^3^ s*^−^*^1^)
*T*: exposure period (s)
*T R*: transmission ratio
*V*: space volume (m^3^)
*V*: viable fraction of RNA copies

## Footnotes

1 https://doi.org/10.1101/2021.11.24.21266807v3

2 A *buffer solution* resists a change in its pH when a small quantity of acid or alkali is added to it

