## Supplemental Materials for "A population framework for predicting the proportion of people infected by the far-field airborne transmission of SARS-CoV-2 indoors"

---

*Keywords:* relative exposure index, airborne, ventilation, aerosols, transmission risk, viral load, COVID-19,

---

*Corresponding author

### 1. Model uncertainties

The output of the REI is a dose value of RNA copies that deposit onto the respiratory tract of a susceptible individual (for brevity we will term this the inhaled dose). The number of viral genome copies (RNA copies) is proportional to the number of viable virion, but the ratio of RNA copies to viable virion is unknown. Whether the deposited virion then leads to an infection in the susceptible individual depends upon the dose response – another unknown quantity, and the susceptibility of the individual to infection.

#### *1.1. Uncertainty in dose response*

There is currently no dose–response curve for SARS-CoV-2, however a number of studies have used a proposed dose curve for SARS-CoV-1, which is a typical coronavirus dose curve [1, 2, 3]. This dose curve was generated from inoculating four groups of transgenic mice (mice genetically modified to express the the human protein that is the receptor for the SARS-CoV-1 virus). The dose response curve was fitted to data from these four groups, in three of which all of the mice became infected and in one group a third of mice become infected. This is a limited data set for curve fitting, although it is sufficient to assume that the dose curve is exponential rather than a Beta-Poisson distribution. It should be noted that the dose response of humans may vary significantly from that of transgenic mice.

Dose curves are fitted to low PFU but there is usually limited data at very low doses. Therefore, the dose-response relationship is highly uncertain at low doses and infection probabilities. This is important because, when considering a large population, even very low probabilities of infection could lead to a significant number of infected people – and it may well be an overestimation if the dose-response curve at very low level of virus is not representative.

In another study Schijven *et al.* determined a model that 1440 RNA copies was required to lead to an infection, drawing assumptions from the proportion of isolated cultured SARS-CoV-2 (ie not collected from patient swabs) needed to infect a cell culture line to calculate a PFU and then deriving an infectious dose from a dose curve for human coronavirus 229E of 1,440 RNA copies [4]. Uncertainties are that SARS-CoV-2 isolate unlikely to be comparable to SARS-CoV-2 collected from patient samples eg swabs. The response of a cell culture is unlikely to be comparable to a respiratory tract, mucosal membranes and innate immunity of a human. The dose curve for 229E may also be different to that for SARS-CoV-2.

There are challenges and uncertainties in the assumptions used to generate infective dose-response curves for SARS-CoV-2 and as these uncertainties are not easily measurable, there will be unknown uncertainties in calculating the probability of infection using such assumptions.

##### *1.2. Uncertainty in viral load*

It has been well established that the viral load of an infector increases from the date of infection and is highest just before or at the onset of symptoms, and as the disease progresses the viral load begins to reduce (within the first week of symptom onset) [5, 6]. Viral load at any stage of infection also varies between individuals, which increases the uncertainty in this value [7, 8, 9, 10, 11, 12]. Some studies use reported cycle threshold values from real time reverse transcription quantitative polymerase chain reaction (RT qPCR) nasopharyngeal (NP) swabs to infer the viral load in respiratory fluid, however this method assumes a direct correlation between the swab viral load and the respiratory fluid viral load [13, 14]. RT qPCR is semi-quantitative in that the number of cycles required to provide a positive signal for SARS-CoV-2 genome is proportional to the starting amount of viral genome in the sample. The greater the number of amplification cycles required, the lower the starting amount of viral genome. A calibrated standard curve can then be used to estimate the starting amount of viral genomic material. However, the standard curve varies between test assays and different RT qPCR thermal cyclers. This method assumes a complete doubling of genetic material at each cycle, and because of the logarithmic relationship, the errors in calculating the starting genomic material for low cycle counts are orders of magnitude higher than those with high cycle counts. Additionally, the estimated concentration of genomic material per unit volume is related to the amount of genomic material in the buffer solution used in the assay, not necessarily the amount in the patients respiratory fluid if data is from NP swabs. The amount of genomic material added to the buffer solution is dependent on not only the viral load of the patient, but also the quality of NP sample collection, which is highly variable. Therefore, it is not possible to determine absolute values of the viral load in patient's respiratory fluid using this method, although it is indicative of a range of variability – much of which is likely to be proportional to the viral load of the individual at the time the sample was collected. While it is somewhat correlated, recent data suggests that the viral load of NP swabs may not reflect the amount of infectious material present [10]. However, it is important to note that there are wide variations in the measured genomic material in NP swabs and that viral load in respiratory fluid is likely to vary over several orders of magnitude, although absolute values and proportions are not determinable with current data.

The RT-qPCR process also only amplifies a small section of viral genome and is representative of viral genomic material in the original sample. Some of this genomic material will be fragments, and therefore quantities of genomic material are not representative of the number of viable virions in the original sample, although likely to be proportional to, and there is some evidence in the literature to suggest there is some correlation between Ct values and infectious virus [6]. Additionally, one study of the influenza virus showed that the viral load in NP swabs was not a significant predictor of aerosol shedding [15]. In other studies the SARS-CoV-2 viral load of saliva has been estimated using qRT-PCR that also show wide variability of several orders of magnitude, although it is unknown if the saliva viral load is the same as the viral concentration in the fluid of the respiratory tract[16].

###### *1.2.1. Viral load in aerosols*

If the viral load in respiratory fluid could be determined it is currently unclear whether the viral concentration in respiratory aerosols and droplets is uniformly distributed. Some studies suggest that the amount of virion in smaller aerosols ($< 1\,\mu$m) is higher than would be expected given the viral concentration in the respiratory fluid [17, 18] and that there may be more genomic material in the smallest aerosols [19]. There is also high variability in the total volume of aerosols generated per unit volume of exhaled breath between individuals, which is especially true for breathing and is dependent upon the respiratory activity and respiratory capacity (e.g. talking, singing) [20, 21, 22, 23]. A recent study from Coleman *et al.* [19] has detected SARS-CoV-2 genomic material in expirated aerosols from *some* Covid patients, although 41% percent of patients exhaled no detectable genomic material. Singing and talking generally produced more genomic material than breathing, but there was large variability between patients. This suggests that respiratory activities that have previously been shown to increase aerosol mass, also increase the amount of viral genomic material, although in this study the viral concentration in aerosols cannot be determined because the mass of aerosols generated was not measured. It also shows that the variability in the amount of genomic material measured in expirated aerosols is consistent with the variability of viral loads as measured by swabs and saliva [19]. Similarly Adenaiye *et al.* have also detected genomic material in aerosols from patients infected with SARS-CoV-2 providing a sampled of exhaled air with some talking and singing. Genomic material was most likely to be detected in exhaled aerosols when the viral load of saliva or Mid-turbinate swabs (MTS) was high ($> 10^8 RNA\, copies$ and $> 10^6$ for MTS and saliva samples respectively). Additionally they were also able to culture viable virus from $< 2\%$ of fine aerosol samples (although one culture positive sample was from a fine aerosol sample which has a less than Limit Of Detection amount of genomic material as measured by RT-PCR, so could be an artefact). Providing some evidence to support the epidemiological evidence that viable virus can exist in exhaled aerosols [24].

Buonanno *et al.*, although noting that there are no values available in the literature, propose a method to convert viral load to quanta emission rate (where a quantum is defined as the dose of airborne droplet nuclei required to cause infection in 63% of susceptible people) using a value for PFU per quanta derived from Watanabe *et al.*, see Section 1.1, and RNA copies per PFU from Fear *et al.* - values derived from stock SARS-CoV-2 created from Vero E6 cells, values which may well not reflect the quanta emission rate in an infected person [25, 1, 26]. There is likely much uncertainty in this method and how representative it is of infector viral emission rates.

##### *1.3. Estimating viral emission from viral load*

Although we have a range of viral loads for infectors in RNA copies per ml, estimating how that relates to the emisison of rnac per unit time is challenging due to the uncertainties listed, however, if we assume that the RNA copies concentration is constant in aerosols and NP swabs we can use the assumptions of Jones *et al.* [27] to convert a NP viral load into a viral shedding rate. This methodology is derived from the aerosol volume distribution of different respiratory activities from Morawskwa *et al.* and is similar to that used by Lelieveld *et al.* [28, 23]. Table 1 shows that for a viral load of $10^7$RNA copies per ml this would assume the RNA copies shedding per hour, and for comparison median values from Coleman *et al.* study are given, in which the measured collected RNA copies were from Covid patients with a median Ct of 16 from the patient's diagnostic sample [19].

Table 1: Estimated RNA copies shedding rates from an infector with a viral load of $10^7$RNA copies per ml compared to measured RNA copies shedding rates from patients with a median Ct of 16 as measured by Coleman *et al.* *value calculated from breathing and talking values

| | estimated $RNA\,copies h^{-1}$ | measured median $RNA\,copies h^{-1}$ |
|---|---|---|
| Breathing | 203 | 127 |
| Voiced counting (talking) | 967 | 1912 |
| Vocalisation (singing) | 6198 | 2856 |
| Breathing:talking 25:75 | 394 | 573* |

Additionally a recent pre-print from Adenaiye *et al.* has also measured viral genome in patients, infected with SARS-CoV-2 alpha variant, breathing with some talking in coarse ( $5\mu m$) and fine ($\leq 5\mu m$) aerosols with a total geometric mean of 1440 $RNA\,copies h^{-1}$ (with a maximum of 3 x $10^5$ $RNA\,copies h^{-1}$) [24]. Although this is more than the estimated values in Table 1, the viral load as measured by genome copies from Mid-turbinate swabs (MTS) was generally orders of magnitude higher than $10^7$.

In the measured data we don't know the relationship between the PCR cycle threshold and the patient viral load in $RNA\,copies/ml$, however the calculated shedding rate of viral genome for a viral load of $10^7$RNA copies per ml is a reasonable fit to the Coleman *et al.* and Adenaiye *et al.* data.

Other studies have suggested that genome emission rates of patients could be of the order of $10^6 RNA\,copies h^{-1}$. Miller *et al.* derived this value from RNA copies measured in small hospital rooms containing Covid patients (here the air sampling equipment is located quite close to the patient and some observation that patients face turned to face collector for some samples) [29,

14]. Whilst Ma *et al.* collected viral genome in exhaled breath of patients that would suggest patients exhaled in the region of 7x$10^4$ to 7x$10^6$ RNA copies per hour, though in this study the collection mechanism involved exhaling into a small straw–like tube for 5 minutes, which could also become contaminated with viral laden saliva, thus over estimating the viral load of the exhaled breath [30]. Although these exhaled rates of viral genome are much greater than those collected by Coleman *et al.*, Miller *et al.* notes that suggests that around 1 : 1000 genome copies are likely to be infectious virion [31, 14]. Adenaiye *et al.* suggest that from MTS there is around 1 : $10^4$ viable virus per measured genome copies[24]. For this study we have made the assumption that all genome copies are viable virion, which either over-estimates the likely infectiousness if using the Coleman *et al.* data, or is similar to the Miller *et al.* assumptions if the viable virion shedding rate is in the order of 1000 virion per hour.

For the proportion of persons infected analysis, the inhaled dose is calculated for all viral loads, it should be noted that the calculated RNA copies shedding rate is assumed to scale linearly with viral load per ml of respiratory fluids, such that a viral load of $10^8 RNA\,copies/ml$ would have ten fold greater RNA copies shedding rates per hour. For comparison, given a viral shedding of 394 RNA copies per hour (assumed for a viral load of $10^7 RNA\,copies/ml$) would lead to an individual inhaled dose of around 2.2 and 0.2 RNA copies for the Small Office and Big Office scenarios respectively.

###### *1.3.1. Comparison of viral emission from literature*

Extrapolating data for viral shedding rates from the literature is challenging as often the estimated doses and the probability of infection do not align with epidemiological evidence. Chen *et al.* suggests that the upper limit for the total virion shedding rate for moderate talking is 6000 virions per hour. This includes droplets up to 100 $\mu$m and so we assume the evaporation and suspension of all these droplets in the air, although it is unlikely. Given a 600 $m^3$ 20 person office at $10\,l\,s^{-1}$ per person with an infected person shedding at 100 virions per minute we would expect $< 10$ virions to deposit in the respiratory tract of a susceptible person over an 8 hour day, which, from the DeDiego *et al.* SARS-CoV-1 dose curve, is unlikely to lead to an infection. This suggests that if the upper limit of viral shedding is unlikely to result in an infection, than the more likely lower viral shedding rates will be even more unlikely to give rise to infection [32, 12].

Using the shedding rate of 6000 virions per hour for the Skagit choir (Miller *et al.* [14] suggest shedding at 1000 virions per hour would be a reasonable estimate for this scenario) we expect a susceptible person to have about 7 virions deposit in their respiratory tract over a 2.5 hour practice period. Using the SARS-CoV-1 dose curve, this gives a probability of infection of 0.02. Given that the secondary attack rate at the Skagit choir was over 85%, this would suggest that either the $k$ value in the SARS-CoV-2 dose curve (see Equation 2) is much smaller than that predicted for SARS-CoV-1 or these models have used assumptions that have under estimated the virion shedding rate, even for the high viral emitter considered here.

Alternatively, the quanta metric could be used because it captures the effects of virion shedding and the dose curve by associating secondary transmission in a particular transmission event. The quanta for the Skagit event is calculated by Miller *et al.*, where the dose is likely to be $1.19 \pm 0.48$ quanta.

Using the Wells-Riley model gives a probability of transmission of between 0.51 and 0.81. This suggests that the quanta method is a better fit in this Skagit choir scenario, although this is to be expected because the quanta emission rate is exclusively derived from the number of secondary transmission events that occurred during the scenario.

If we use the Skagit quanta emission rate in another scenario, say a UK junior school classroom described by Jones *et al.* [27], then it is possible to conclude the following: the emission rate for singing is $970 \pm 390\,\mathrm{q\,h^{-1}}$ but assume a 30-fold reduction for aerosol emission when breathing and assume a child breath rate ($q_{sus}$) of $0.44\,\mathrm{m^3\,h^{-1}}$, then the dose over a 7 hour exposure period is $0.77 \pm 0.31$ quanta, giving a probability of infection between 0.37 and 0.66. Although transmission events do occur in school classrooms, there isn't evidence to suggest such high rates of secondary far-field transmission occur regularly. Secondary attack rates (for all routes of transmission) amongst primary pupils have been recorded at less than 1% [33] (although more recent observations on infection rates amongst UK school age children in Autumn 2021 suggests secondary attack rates are likely to be higher than this for the Delta variant, however, still not at probability of infection between 0.37 and 0.66 [34]). This suggests that the quanta emission rate (as estimated in the Skagit Choir scenario) is either extremely unlikely or it scenario-specific and so it is inappropriate to use a quanta emission rate determined from a single scenario and apply to another. Sze *et al.* also covers uncertainties in the use of quanta models [35].

#### 2. Results and discussion: Effect of varying model assumptions on *PPI*

In addition to the results reported in the main paper, here we report the effect of various model assumptions on the *PPI* and *TR*. All standard scenario inputs are given in Tables 2 and 3.

Table 2: Scenario inputs and calculations of individual risk.

| | Big Office Reference | Small Office Comparator |
|---|---|---|
| Number of occupants, $N$ | 50 | 5 |
| Space Volume, $V$ ($m^3$) | 1500 | 150 |
| *Per capita* volume, $V\,N^{-1}$ ($m^3$ per person) | 30 | |
| Air flow rate, $\psi V$ ($l\,s^{-1}$) | 500 | 50 |
| Air change rate, $\psi$ ($h^{-1}$) | 1.2 | |
| Removal rate, $\phi$ ($h^{-1}$) | 2.26 | |
| Equivalent ventilation rate , $\phi V$ ($l\,s^{-1}$) | 942 | 94.2 |
| Exposure time, $T$ (h) | 8 | |
| Dose constant, $k$ [32] | 410 | |
| Respiratory tract absorption fraction, $K$ | 0.55 | |
| Viable fraction, $v$ (%) | 100 | |
| Viral load (RNA copies per ml) [36] | $10^7$ | |
| Respiratory activity, *breathing:talking* (%) | 72:25 | |
| Volumetric ratio of exhaled droplets to exhaled air, $V^*_{drop}$ | $5.05 \times 10^{-13}$ | |
| Respiratory fluid density ($ml\,m^{-3}$) | $1.25^8$ | |
| Respiratory rate, $q_{resp}$ ($m^3\,h^{-1}$) | 0.56 | |
| Viral load, $L$ () | $10^7$ | |
| Viral emission rate, $G$ (RNA copies per hour) | 394 | |
| Community infection rate, $C$ | 1:100 | |
| Dose, $D$ (viable virions inhaled) | 0.245 | 2.450 |
| REI | 1 | 10 |

All values converted to SI units before application.

Table 3: Scenario inputs and calculations of population risk.

| | Big Office Reference | Small Office Comparator |
|---|---|---|
| Viral load [36] () | N(7,1.4) | |
| $P(R)$ (%) | 0.062 | 0.620 |
| $P(I=0)$ (%) | 61 | 95 |
| $P(0 < I < N)$ (%) | 39 | 5 |
| $\bar{I}$ | 1.27 | 1.02 |
| $P(S)$ (%) | 39 | 5 |
| $PPI$ (%) | 1.59 | 0.43 |
| $TR$ | 0.27 | |

N, normal($\mu$,$\sigma$)

All values converted to SI units before application.

##### 2.1. *Dose curve constant k*

Figure 1 shows that when the dose k values are low (< 50) then the $PPI$ begins to increase rapidly as lower doses are required to result in a significant proportion of susceptibles in a scenario population becoming infected. The rate of increase in $PPI$ is greater in Big Office due to the larger population of susceptibles. This results demonstrate that the dynamics of the dose response are important in understanding the $PPI$ and more work is required to better understand these characteristics for SARS-CoV-2. Epidemiological evidence can provide some illumination as to what bounds values of $k$ with respect to measured far-field transmission rates in indoor scenarios.

##### 2.2. *Virion viability*

In our study, we take the conservative assumption that all viral genome copies (RNA copies) in an inhaled dose are viable virions. The actual proportion it more likely to be orders of magnitude lower, with estimates in the

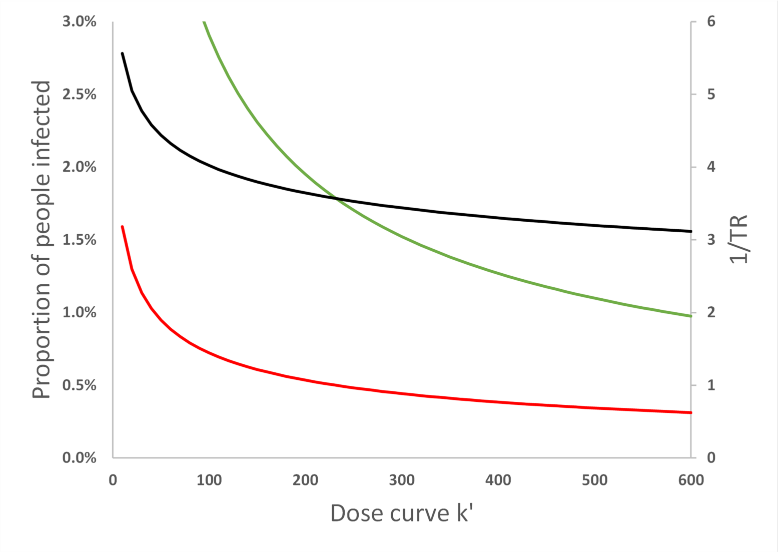


Figure 1: The effect of increasing the dose curve constant, $k$, on the Big Office$PPI$ (green), Small Office$PPI$ (red) and the $TR$ (black). As $k$ increases the size of the inhaled dose needed to give an equivalent probability of infection increases. All values are illustrative.

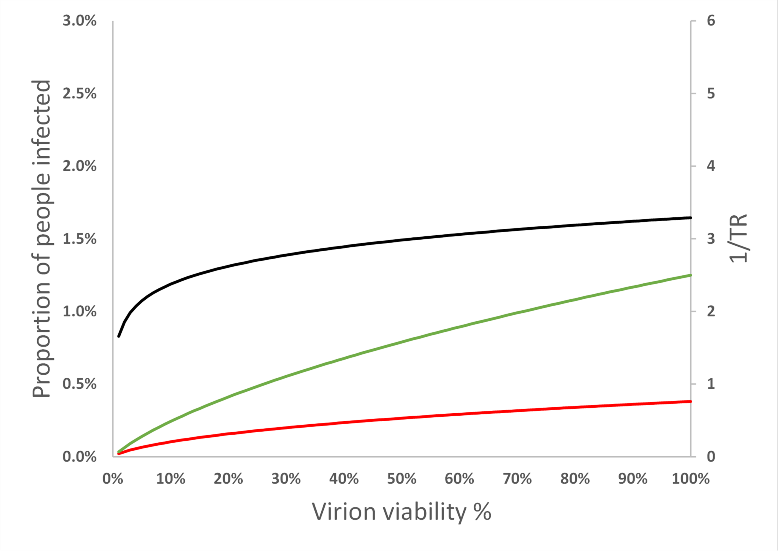


Figure 2: The effect of increasing the virion viability on the Big Office$PPI$ (green), Small Office$PPI$ (red) and the $TR$ (black). As the proportion of RNA copies in the inhaled dose that are viable of virions increase, the probability of infection increases. All values are illustrative.

literature of a range of 1 : 100 to 1 : 10000 of viable virions to RNA copies. Miller *et al.* suggests that around 1 : 1000 genome copies are likely to be infectious virion whilst Adenaiye *et al.* suggest that from mid-turbinate swabs there is around 1 : $10^4$ viable virus per measured genome copies[14, 24].

Figure 2 shows the *PPI* in both Big Office and Small Office reduces ss the proportion of viable virions decreases. Whilst the *TR* decreases, the ratio of Small Office to Big Office remains above 2, but the absolute values of *PPI* become very low as virion viability < 1%, suggesting that far-field transmission, given the assumptions in Tables 2 and 3, is very unlikely if virion viability is low.

##### *2.3. Viral Load of infected*

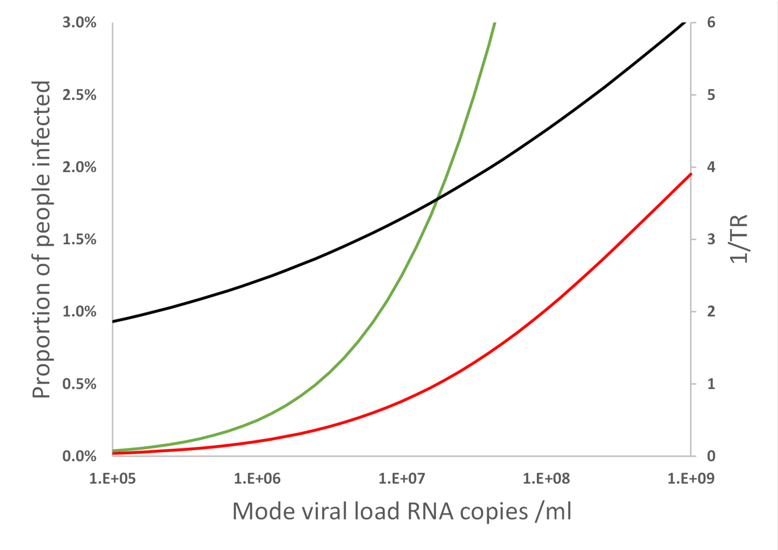


Figure 3: The effect of increasing the mode viral load of the infected population on the Big Office *PPI* (green), Small Office *PPI* (red) and the *TR* (black). As the mode viral load of the infectors increases, the emission rate of RNA copies increases, resulting in an increase in dose and *PPI*. All values are illustrative.

The inhaled dose is also a function of the viral load distribution within the infected population. We assume that the log 10 viral load in RNA copies per ml is normally distributed with a mean of $\mu = 7$ and a standard deviation of $\sigma = 1.4$. Figure 3 shows the change in the probability of transmission and the $TR$ when the mean log value of the distribution is varied between 5 and 9. When the mean is low, the probability of one or more infectors having a sufficiently high emission rate to lead to the inhaltion of an infective dose is very low, ie when $\mu = 5$. Conversely, increasing the probability of the infectors having a high viral load (by increasing $\mu$) rapidly increases the probability of transmission in both scenarios, and an increase in the $TR$.

##### 2.4. Space Volume per person

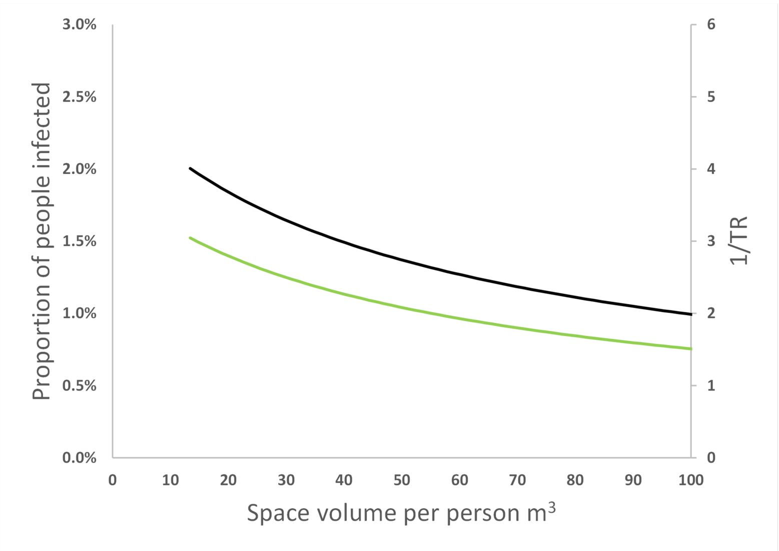


Figure 4: The effect of increasing the *per capita* space volume, $V$, in the Big Office on the $PPI$ (green) and the $TR$ (black) when the *per capita* space volume in the Small Office is constant. All values are illustrative.

Figure 4 shows that increasing the *per capita* space volume in the Big Office when the *per capita* space volume in the Small Office, while maintaining a constant *per capita* ventilation in both spaces has a similar effect to increasing the *per capita* ventilation. This is because the dose is inversely proportional to volume. Furthermore, the product of the space volume and the total removal rate, $\phi V$, is proportional to the concentration of the virus in the air and, therefore, the dose. The *per capita* ventilation rate is constant in both spaces and so the air change rate in the Big Office decreases as its volume increases. However, this reduction is offset by the surface deposition and biological decay rates, which remain constant and have a greater effect on the value of the equivalent ventilation rate, $\psi V$, as the space volume increases.

Equation 1 assumes a steady-state concentration of the virus has been reached based on the assumption that the exposure time, $T$, is significant. However, the time taken to reach the steady-state concentration in large spaces may be significant and affects the dose over shorter exposure periods. This is an example of the *reservoir effect*, the ability of indoor air to act as a fresh-air reservoir and absorb the impact of contaminant emissions. The greater the space volume, the greater the effect. These factors highlight the benefits of increasing the *per capita* space volume.

##### *2.5. Exposure Time*

Increasing exposure time when an infected person is present in the space for a significant period of time the exponent of Equation 1 becomes relatively small so that $e^{-\phi T} \to 0$ and the inhaled dose is approximately proportional to the exposure time, however, the effect of the dose curve relationship means that $PPI$ is not directly proportional to exposure time. Reducing the exposure time from 12 to 4 hours will reduce the probability of an inhaled dose

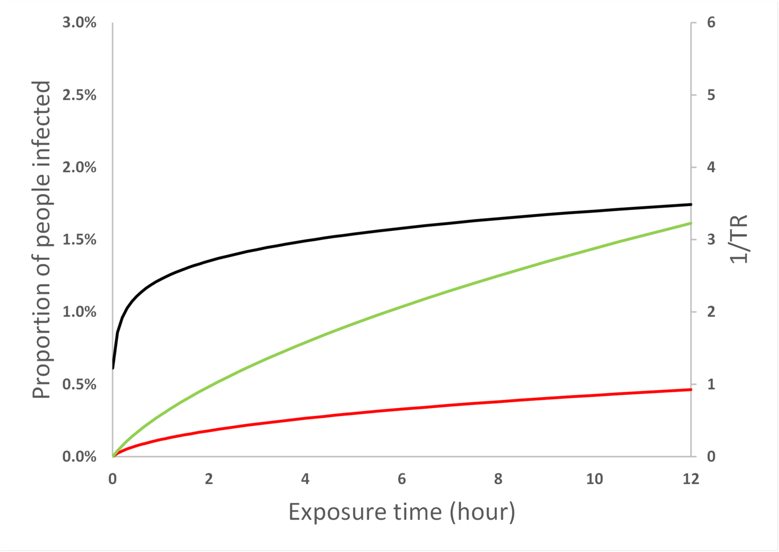


Figure 5: The effect of increasing the exposure time on the Big Office $PPI$ (green), Small Office $PPI$ (red) and the $TR$ (black). As the exposure time increases, the dose increases, resulting in an increase in $PPI$. All values are illustrative.

leading to infection from relatively low viral load infectors, but will have less effect on the higher viral load infectors. It is only when exposure times become very short that the $PPI$ reduces rapidly due to the reduced probability of even the higher viral load infectors delivering a dose likley to lead to infection, (as this study assumes steady state, this rate of reduction will be more pronounced when considering the reservoir effect) this is consistent with the findings of Miller *et al.* in the requirement for reduced exposure time, as well as improved ventilation to significantly reduce the risk of transmission in the case of the Skagit choir superspreading event [14].

#### 3. Rapid Antigen Testing

Lateral flow testing uses a rapid lateral flow device (LFD) based on colloidal gold immunochromatography designed to detect the presence of SARS-

CoV-2 nucleocapsid antigens in nasopharyngeal swabs. These tests are not as sensitive as PCR tests in detecting the presence of SARS-CoV-2, but they have been demonstrated to have a good ability at detecting higher viral loads in infectors [37, 38]. Because the results of our analysis demonstrates that the higher viral emissions are responsible for the greater $PPI$ we consider the effect of a scenario where widespread adoption of LFD use is successful in identifying individuals with high viral load and removing them from the scenario. The distribution of log 10 viral loads of infectors is assumed to be normally distributed with a mean of $\mu = 7$ and a standard deviation of $\sigma = 1.4$, then the proportion of individuals with viral loads greater than $10^9$RNA copies per ml is about 9%. We can assume a proportion of these are removed from the Small Office and Big Office scenarios and consider the effects on the $PPI$ given the assumptions in Table 2. The probability of viral load, $P(L)$ when $VL > 10^9 RNA\, copies/ml$ is multiplied by $1 - LFD_{effectiveness}$ where the effectiveness is assumed to be 70%, Figure 6. These results show that LFD could be an effective measure to reduce the $PPI$, reducing both the absolute $PPI$ in Big Office (from 1.59 to 0.60) and Small Office(from 0.43 to 0.22), as well as the $TR$. However, with a $C$ of $1 : 100$, persons with a viral load greater than $10^9 RNA\, copies/ml$ represent around 0.09% of the total population, so although LFD could be an effective method of removing the highest viral loads from a scenario, a lot of lateral flow tests need to be conducted to capture every high viral load infector.

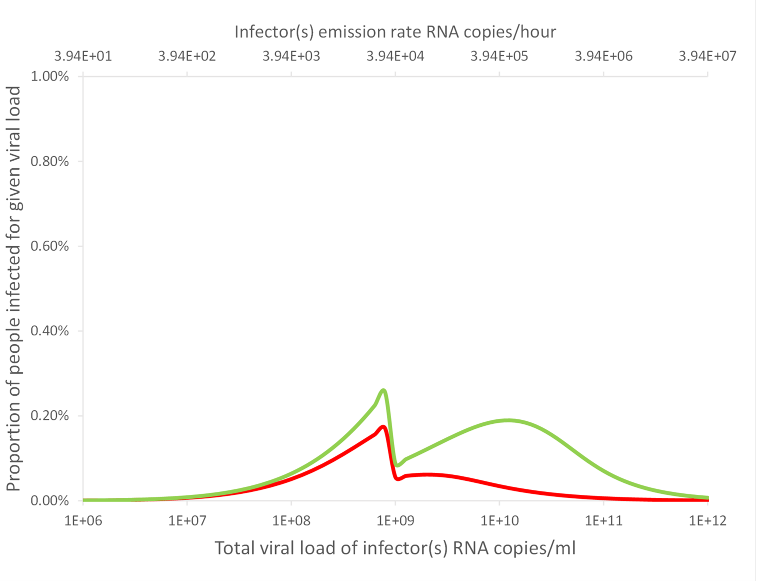


Figure 6: An indication of the relationship between the proportion of a population infected for a particular viral load when the community infection rate is $C = 1\%$ and where LFD that are 70% effective at removing infectors with viral loads greater than $10^9$ RNA copies per ml. The area under the curve represents the total proportion of people infected for the Small Office (red) and the Big Office (green). All values are illustrative.

#### 4. Alternative assumptions, lower virion viability and higher mode viral load

As detailed previously, the assumptions used in the main paper with respect to how RNA copies represent viable virions is highly conservative, and so below we use a more realistic value of 1% RNA copies as representing viable virions. Additionally we also consider that the mode value for viral load is $10^8$ RNA copies per ml, to represent a potential increase in viral load that could be the result of a variant of SARS-CoV-2, see Tables 4 and 5.

Figure 7 shows how the reduction in virion viability shifts the Big Office and Small Office dose curves to the right of the graph as greater viral emission is required to result in a dose of viable virion likely to give rise to an infection. The viral load and the probability that a single has that viral load, $P(L)$, is also shifted to the right. The dashed vertical lines show the viral load required to give a 50% probability that the dose will lead to an infection for each scenario, $P(R) = 50\%$. The area under the blue curve to the right of each vertical line is the probability that the viral load of the infected person leads to $P(R) \geq 50\%$. The probability is much smaller for the Big Office, which has the lower REI. This probability that an infected person has a viral load that leads to $P(R) \geq 50\%$ is small, suggesting that the most likely outcome is $P(R) \leq 50\%$.

The effect of varying assumptions on the shape of the plotted $PPI$ curves have been described in detail in the main paper and above 2. In Figures 8 and 9 the affect on $PPI$ and $TR$ is shown for the assumptions made in the main paper (on the left) with the higher modal viral load and lower virion viability (on the right). The effect on the absolute values is pronounced due to the reduced virion viability assumption, and given these assumptions the $PPI$ is very low, suggesting that far field transmission is likely to be rare and efforts taken to minimise $TR$ should consider the absolute improvements in $PPI$ when assessing the benefits in $TR$ reduction compared to the costs of, for example, the increased energy use needed to increase ventilation above current guidance. It is important to note how changes in assumptions needed to estimate viral emission rates have large impacts on absolute values of $PPI$ and thus the uncertainty in these assumptions needs to be considered when interpreting comparisons.

Table 4: Scenario inputs and calculations of individual risk.

| | Big Office Reference | Small Office Comparator |
|---|---|---|
| Number of occupants, $N$ | 50 | 5 |
| Space Volume, $V$ ($m^3$) | 1500 | 150 |
| *Per capita* volume, $V\,N^{-1}$ ($m^3$ per person) | 30 | 30 |
| Air flow rate, $\psi V$ ($l\,s^{-1}$) | 500 | 50 |
| Air change rate, $\psi$ ($h^{-1}$) | 1.2 | 1.2 |
| Removal rate, $\phi$ ($h^{-1}$) | 2.26 | 2.26 |
| Equivalent ventilation rate , $\phi V$ ($l\,s^{-1}$) | 942 | 94.2 |
| Exposure time, $T$ (h) | 8 | 8 |
| Dose constant, $k$ [32] | 410 | 410 |
| Viable fraction, $v$ (%) | **1** | **1** |
| Viral load (RNA copies per ml) [36] | $\mathbf{10^8}$ | $\mathbf{10^8}$ |
| Respiratory activity, *breathing:talking* (%) | 75:25 | 75:25 |
| Viral emission rate, $G$ (RNA copies per hour) | 394 | 394 |
| Respiratory rate, $q_{sus}$ ($m^3h^{-1}$) | 0.56 | 0.56 |
| Community infection rate, $C$ | 1:100 | 1:100 |
| Dose, $D$ (viable virions inhaled) | **0.002** | **0.025** |
| REI | 1 | 10 |

All values converted to SI units before application.

For this analysis we make the assumption that 1% of genome copies

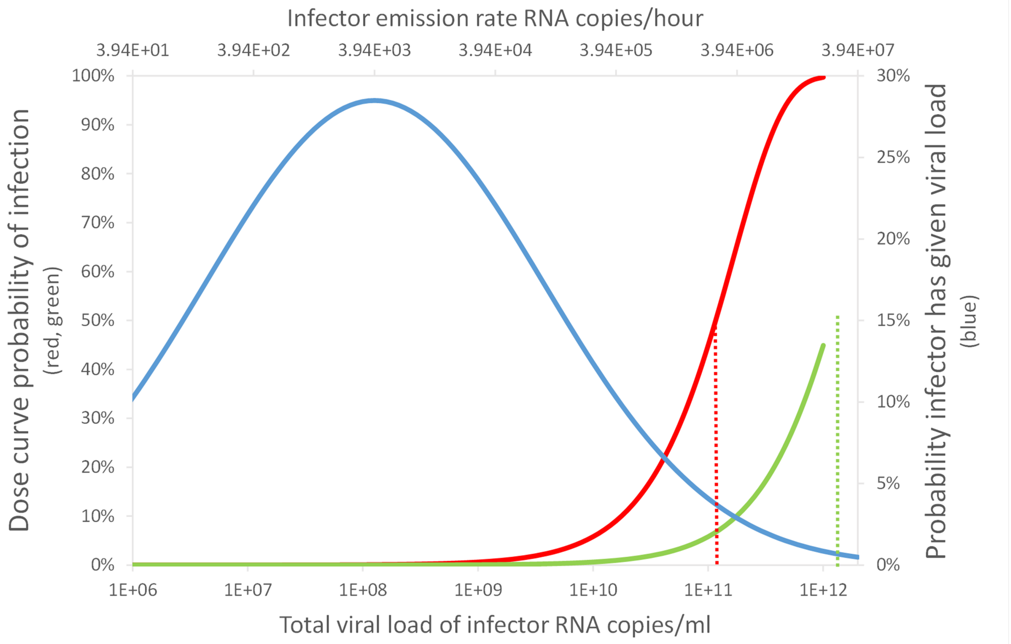


Figure 7: An indication of the relationship between the viral load, $L$, and the consequent probability of infection, $P(R)$, in the Big Office (green) and Small Office (red) for a susceptible occupant, and the probability of a single infected person having a viral load, $P(L)$, (blue). Dashed vertical lines indicate the viral load required for $P(R) = 50\%$.

Table 5: Scenario inputs and calculations of population risk.

| | Big Office Reference | Small Office Comparator |
|---|---|---|
| Viral load [36] (RNA copies per ml) | **LN(**$2.7 \times 10^{10}$,$3.6 \times 10^{11}$**)** | |
| $P(R)$ (%) | 0.062 | 0.620 |
| $P(I = 0)$ (%) | 61 | 95 |
| $P(0 < I < N)$ (%) | 39 | 5 |
| $\bar{I}$ | 1.27 | 1.02 |
| $P(S)$ (%) | 39 | 5 |
| $PPI$ (%) | **0.241** | **0.119** |
| $TR$ | 0.49 | |

LN, log-normal($\mu$,$\sigma$)

All values converted to SI units before application.

(RNA copies) represent viable virions, which is a more realistic magnitude to assume given the literature

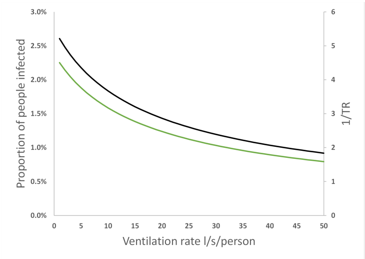


(a) Big Office Ventilation

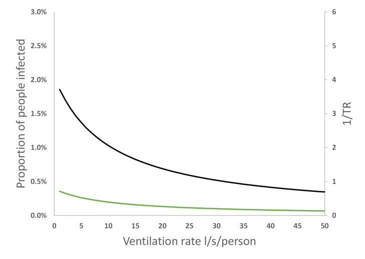


(b) Big Office Ventilation

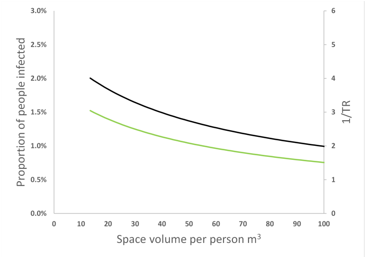


(c) Big Office Space volume

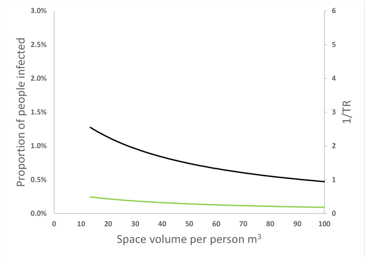


(d) Big Office Space volume

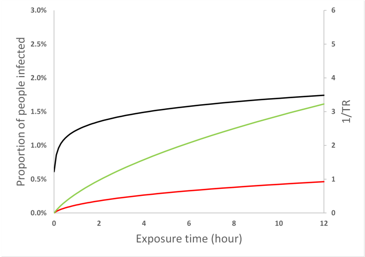


(e) Exposure time

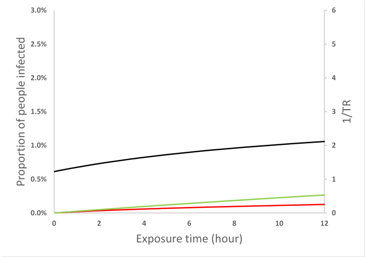


(f) Exposure time

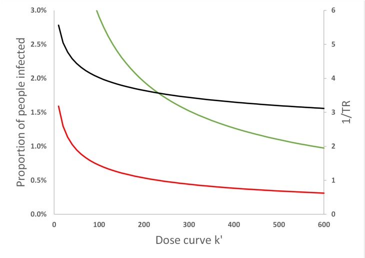


(g) Dose constant $k$

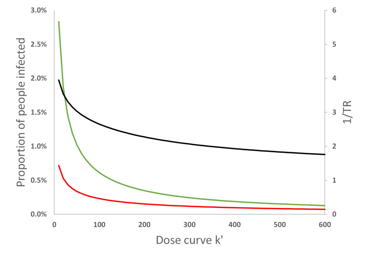


(h) Dose constant $k$

Figure 8: The effect of modulating assumption values on the Big Office $PPI$ (green), Small Office $PPI$ (red) and the $TR$ (black). Left hand images the modal viral load assumed to be $10^7$ RNA copies per ml and virion viability 100%, right hand images the modal viral load assumed to be $10^8$ RNA copies per ml and virion viability 1%. All values are illustrative.

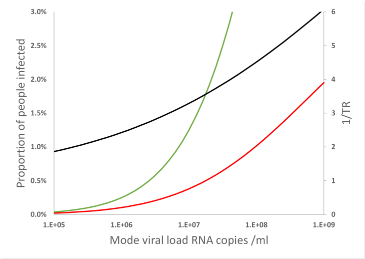


(a) Modal viral load

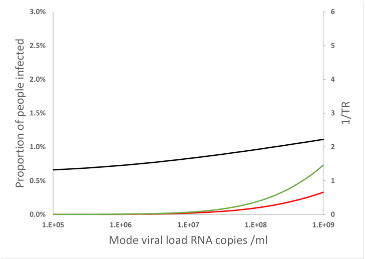


(b) Modal viral load

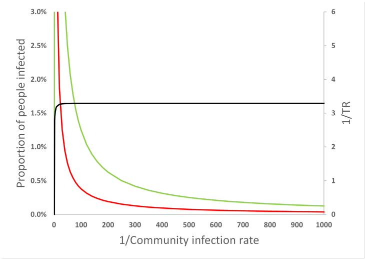


(c) Community infection rate

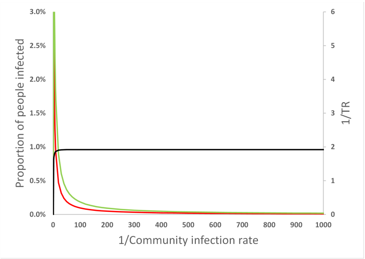


(d) Community infection rate

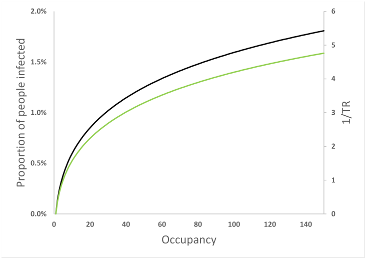


(e) Occupancy (*per capita* space volume and ventilation preserved)

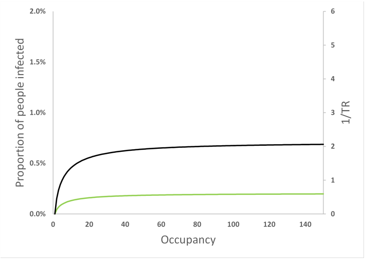


(f) Occupancy (*per capita* space volume and ventilation preserved)

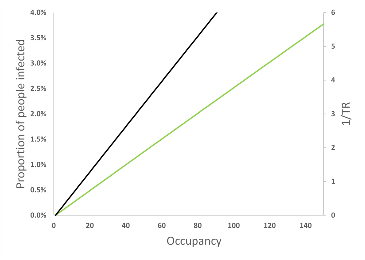


(g) Occupancy (Fixed volume and ventilation, 1500m$^3$ and 500l per s, respectively)

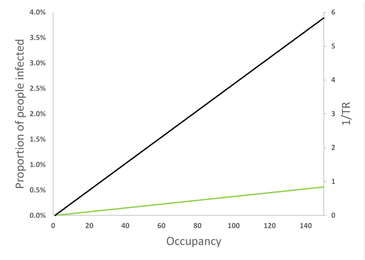


(h) Occupancy (Fixed volume and ventilation, 1500m$^3$ and 500l per s, respectively)

Figure 9: The effect of modulating assumption values on the Big Office *PPI* (green), Small Office *PPI* (red) and the *TR* (black). Left hand images the modal viral load assumed to be $10^7$ RNA copies per ml and virion viability 100%, right hand images the modal viral load assumed to be $10^8$ RNA copies per ml and virion viability 1%. All values are illustrative.

#### 5. Equations

See main text for explanations of the equations.

##### 5.1. Inhaled Dose

$$D \simeq \frac{K\, q_{sus}\, G\, T\, v}{\phi\, V} \tag{1}$$

##### 5.2. Dose response curve

$$P(R) = 1 - e^{-D/k} \tag{2}$$

#### Acknowledgements

The authors acknowledge the Engineering and Physical Sciences Research Council (EP/W002779/1) who financially supported this work. They are also grateful to Constanza Molina for her comments on this paper.

[34] ONS, ONS Coronavirus (COVID-19) Infection Survey.

URL https://www.ons.gov.uk/peoplepopulationandcommunity/healthandsocialcare/conditionsanddiseases/bulletins/coronaviruscovid19infectionsurveypilot/previousReleases

[35] G. N. Sze To, C. Y. H. Chao, Review and comparison between the wells–riley and dose-response approaches to risk assessment of infectious respiratory diseases, Indoor air 20 (1) (2010) 2–16. doi:10.1111/j.1600-0668.2009.00621.x.

[36] P. Z. Chen, N. Bobrovitz, Z. Premji, M. Koopmans, D. N. Fisman, F. X. Gu, Sars-cov-2 shedding dynamics across the respiratory tract, sex, and disease severity for adult and pediatric covid-19, eLife 10 (2021) e70458. doi:10.7554/eLife.70458.
URL https://doi.org/10.7554/eLife.70458

[37] J. Ferguson, S. Dunn, A. Best, J. Mirza, B. Percival, M. Mayhew, O. Megram, F. Ashford, T. White, E. Moles-Garcia, L. Crawford, T. Plant, A. Bosworth, M. Kidd, A. Richter, J. Deeks, A. McNally, Validation testing to determine the sensitivity of lateral flow testing for asymptomatic SARSCoV-2 detection in low prevalence settings: Testing frequency and public health messaging is key, PLoS Biology 19 (4) (2021) 1–9. doi:10.1371/journal.pbio.3001216.
URL http://dx.doi.org/10.1371/journal.pbio.3001216

[38] L. Y. W. Lee, S. Rozmanowski, M. Pang, A. Charlett, C. Anderson, G. J. Hughes, M. Barnard, L. Peto, R. Vipond, A. Sienkiewicz, S. Hopkins, J. Bell, D. W. Crook, N. Gent, A. S. Walker, T. E. A. Peto, D. W. Eyre, Severe Acute Respiratory Syndrome Coronavirus

2 (SARS-CoV-2) Infectivity by Viral Load, S Gene Variants and Demographic Factors, and the Utility of Lateral Flow Devices to Prevent Transmission, Clinical Infectious Diseases 6 (2021) 1–32. doi:10.1093/cid/ciab421.
URL https://academic.oup.com/cid/advance-article/doi/10.1093/cid/ciab421/6273394
