## Supplementary material for "A population framework for predicting the proportion of people infected by the far-field airborne transmission of SARS-CoV-2 indoors": Matlab code: mc.m

```
function data = mc
%%%%%%%%%%%%%%%%%%%%%%%%%%%%%%%%%%%%%%%%%%%%%%%%%%%%%%%%%%%%%%%%%%%%%%%%%%%
%
% mc.m
%
% author: Benjamin M. Jones
%
% INPUTS
% none: 
%       
%
% OUTPTUS
% data: a cell aray containing 3 tables.
% data{1}: calcualted parameters for a 50 person space
% data{2}: calcualted parameters for a 5 person space
% data{3}: statistics for reporting
%
% Copyright 2022 Benjamin Michael Jones Licensed under the
% Educational Community License, Version 2.0 (the "License"); you may
% not use this file except in compliance with the License. You may
% obtain a copy of the License at
%     
% http://opensource.org/licenses/ECL-2.0
% 
% Unless required by applicable law or agreed to in writing,
% software distributed under the License is distributed on an "AS IS"
% BASIS, WITHOUT WARRANTIES OR CONDITIONS OF ANY KIND, either express
% or implied. See the License for the specific language governing
% permissions and limitations under the License.
%
% CONDITIONS OF USE: Please note that by downloading and using this file
% you agree to the following terms and conditions.
% i) This software must be cited in any publication (with web link).
% ii) The following papers must be cited in any publication:
%    a) Jones B, et al. Modelling uncertainty in the relative risk of 
%       exposure to the SARS-CoV-2 virus by airborne aerosol transmission 
%       in well mixed indoor air. Building and Environment. 
%       2021;191:107617.
%    b) Idden C, et al. A population framework for predicting the proportion 
%       of people infected by the far-field airborne transmission of 
%       SARS_CoV-2 indoors. 2022; in-review.
% 
%%%%%%%%%%%%%%%%%%%%%%%%%%%%%%%%%%%%%%%%%%%%%%%%%%%%%%%%%%%%%%%%%%%%%%%%%%%

    %% CONSTANTS AND COEFFICIENTS
    % number of occupants
    pop_ratio   = 10;
    N(1,1)      = 50; % reference space
    N(2,1)      = N(1)/pop_ratio; % comparator space
    % number of spaces
    spaces(1,1) = 2e5;
    N_pop       = N(1)*spaces(1); % samples
    spaces(2,1) = N_pop/N(2);
    % space volume
    V(1,1)      = 1500; % m3
    V(2,1)      = V(1)/pop_ratio; % m3
    % DOSE TERMS
    phi         = 2.36;  % removal rate [1/h]
    T           = 8;     % exposure period [h]
    K           = 0.55;  % lung deposition factor [-]
    k           = 410;   % reciprocal of the probability that a single pathogen initiates an infection [-]
    v           = 1;     % viable fraction [-]
    qsus        = 0.558; % breathing rate [m3/hour]
    qinf        = qsus;  % breathing rate [m3/hour]
    C           = 1/100; % community infection rate [-]
    V_s         = 5.05e-13; % Volumetric ratio of exhaled droplets to exhaled air [-]
    DV_G        = K*qsus*T*v/phi; % dose per the product of the volume and emission rate [virions m3.s/RNA copies]
    rfd         = 1.25e8; % respiratory fluid density [ml/m3]
    L_mu        = 7;   % mean viral load [log10 RNA copies per ml]
    L_sigma     = 1.4; % stdev of viral load [log10 RNA copies per ml]
    [P_I,P_S,I_bar,Gt_bar,G_bar,Dt_bar,D_bar,P_Rt_bar,P_R_bar,PPI,ppi] = deal(zeros(2,1)); % initialise output vectors

    %% CALCULATIONS
    % loop through scenario
    for scen = 1:2
        % initialise vectors
        [N_I,N_S,L,G,D,P_R,N_S_I] = deal(zeros(spaces(scen),1));
        % loop through each space
        for space = 1:spaces(scen)
            % number of infected people when susceptible people present
            N_I(space)     = binornd(N(scen),C);
            if N_I(space) == N(scen)
                N_I(space) = 0;
            end
            % number of susceptible and exposed people
            if N_I(space) == 0
               N_S(space)  = 0;
            else
               N_S(space)  = N(scen) - N_I(space);
            end
            % viral load
            L(space)       = sum(10.^(randn([N_I(space),1]).*L_sigma+L_mu)); % [RNA copies per ml]
            % emission rate
            G(space)       = qinf*V_s*rfd.*L(space); % [RNA copies per hour]
            % dose
            D(space)       = DV_G*G(space)/V(scen); % [virions inhaled]
            % probability of infection
            P_R(space)     = 1-exp(-D(space)/k);
            % number of susceptible people infected
            N_S_I(space)   = binornd(N_S(space),P_R(space));
        end
        % collate scenario variables
        data{scen}     = table(N_I,N_S,L,G,D,P_R,N_S_I);
        % calculate means
        P_I(scen)      = sum(N_I>0)/spaces(scen); % Probability of a infected people P(0<I<N) [Eq9]
        P_S(scen)      = sum(N_S)/N_pop;    % Probability of a susceptible people P(S) [Eq10]
        I_bar(scen)    = mean(N_I(N_I>0));  % mean number of infected people when infected people present [Eq12]
        G_bar(scen)    = mean(G(N_I>0));    % mean emission rate
        Gt_bar(scen)   = mean(G);           % mean emission rate
        D_bar(scen)    = mean(D(N_I>0));    % mean dose
        Dt_bar(scen)   = mean(D);           % mean dose
        P_Rt_bar(scen) = mean(P_R);         % mean probability of infection in all spaces
        P_R_bar(scen)  = mean(P_R(N_I>0));  % mean probability of infection when infected people present [Eq11]
        PPI(scen)      = sum(N_S_I)/N_pop;  % proportion of population infected
        ppi(scen)      = P_S(scen)*P_R_bar(scen); % proportion of susceptible population infected [Eq9];
    end

    % transmission ratio [comparator:reference]
    TR = PPI(2)/PPI(1);

    %% POST PRECESS
    data{3}          = table(N,V,spaces,P_I,P_S,I_bar,Gt_bar,G_bar,Dt_bar,D_bar,P_Rt_bar,P_R_bar,PPI,ppi,[TR;TR]);
    data{3}.Properties.VariableNames{end} = 'TR';
    data{3}.P_I      = round(data{3}.P_I*100,2,'significant');
    data{3}.P_S      = round(data{3}.P_S*100,2,'significant');
    data{3}.I_bar    = round(data{3}.I_bar,2,'significant');
    data{3}.Gt_bar   = round(data{3}.Gt_bar,2,'significant');
    data{3}.G_bar    = round(data{3}.G_bar,2,'significant');
    data{3}.Dt_bar   = round(data{3}.Dt_bar,2,'significant');
    data{3}.D_bar    = round(data{3}.D_bar,2,'significant');
    data{3}.P_Rt_bar = round(data{3}.P_Rt_bar*100,2,'significant');
    data{3}.P_R_bar  = round(data{3}.P_R_bar*100,2,'significant');
    data{3}.PPI      = round(data{3}.PPI*100,2,'significant');
    data{3}.ppi      = round(data{3}.ppi*100,2,'significant');
    data{3}.TR       = round(data{3}.TR,2,'significant');
end
```
