## Supplementary material for "A population framework for predicting the proportion of people infected by the far-field airborne transmission of SARS-CoV-2 indoors": Matlab code: plots.m

```
%% GENERAL DATA
font_size  = 10;
font_name  = 'Garamond';
lineWidth  = 0.5;
L_mu        = 7;   % mean viral load [log10 RNA copies per ml]
L_sigma     = 1.4; % stdev of viral load [log10 RNA copies per ml]

%% DOSE CDF
% log_10 = true;
% width = 10;
% height = 8;
% filename = 'dose_cdf';
% h_plot = figure(1);
% 
% D1 = data{1}.D(data{1}.D>0);
% D2 = data{2}.D(data{2}.D>0);
% D1 = data{1}.D;
% D2 = data{2}.D;
% 
% if log_10
%     D1 = log10(D1);
%     D2 = log10(D2);
% end
% 
% p1 = cdfplot(D1);
% hold on;
% p2 = cdfplot(D2);
% 
% grid off;
% set(p1,'Color','k')
% set(p2,'Color','k','LineStyle','--')
% if log_10, xlabel({'Dose (log$_{10}$ virions)'},'Interpreter','latex');
% else xlabel({'Dose (virions)'},'Interpreter','latex'); end
% ylabel('Cumulative Probability','Interpreter','latex');
% title('');
% % axis([0,3,0,1]);
% ax = gca; % Get axis to modify
% if ~log_10, ax.XScale = 'log'; end
% ax.YAxis.FontSize = font_size;
% ax.YAxis.FontName = font_name;
% ax.XAxis.FontSize = font_size;
% ax.XAxis.FontName = font_name;
% if log_10, ax.XLim = [-8 4]; 
% else ax.XLim = [1e-3 1e4]; end
% ax.TickLabelInterpreter = 'latex';
% ax.YLim = [0,1];
% 
% set(gcf, 'PaperPositionMode', 'auto');
% print('-depsc',[filename,'.eps']); 
% set(gcf, 'PaperPositionMode', 'manual');
% set(gcf, 'PaperUnits', 'centimeters');
% set(gcf, 'PaperPosition', [0 0 width height]);
% saveas(h_plot,filename,'png');
% close(h_plot);

%% VIRAL LOAD PDF AND INFECTION RISK
% width = 10;
% height = 8;
% filename = 'load_v_risk';
% h_plot = figure(2);
% hold on;
% % plot the PDF
% L_mu        = 7;   % mean viral load [log10 RNA copies per ml]
% L_sigma     = 1.4; % stdev of viral load [log10 RNA copies per ml]
% pd = makedist('Normal','mu',L_mu,'sigma',L_sigma);
% L = [0.1:0.1:12];
% y = pdf(pd,L);
% % L = log10(L);
% p1 = plot(L,y,'-.k','LineWidth',lineWidth);
% % set(p1,'Color','k')
% xlabel({'Viral Load (log$_{10}$ RNA Copies per ml)'},'Interpreter','latex');
% ylabel('Viral Load Probability Density','Interpreter','latex');
% title('');
% % L = [1e5 1e6:1e4:1e11 1e12];
% L = 10.^L;
% pop_ratio    = 10;
% V(1)         = 1500; % m3
% V(2)         = V(1)/pop_ratio; % m3
% phi          = 2.36;  % removal rate [1/h]
% T            = 8;     % exposure period [h]
% K            = 0.55;  % lung deposition factor [-]
% k            = 410;   % reciprocal of the probability that a single pathogen initiates an infection [-]
% v            = 1;     % viable fraction [-]
% qsus         = 0.558; % breathing rate [m3/hour]
% qinf         = qsus;  % breathing rate [m3/hour]
% V_s          = 5.05e-13; % Volumetric ratio of exhaled droplets to exhaled air [-]
% DV_G         = K*qsus*T*v/phi; % dose per the product of the volume and emission rate [virions m3.s/RNA copies]
% rfd          = 125e6; % respiratory fluid density [ml/m3]
% G            = qinf*V_s*rfd.*L; % emission rate [RNA copies per hour]
% D1           = (DV_G/V(1)).*G;   % dose [virions inhaled]
% P_R1         = 1-exp(-D1./k);    % probability of infection
% D2           = (DV_G/V(2)).*G;   % dose [virions inhaled]
% P_R2         = 1-exp(-D2./k);    % probability of infection
% yyaxis right;
% L = log10(L);
% plot(L,P_R1,'k','LineWidth', lineWidth);
% plot(L,P_R2,'--k','LineWidth', lineWidth);
% L50 = log10(-log(0.5)*k.*V./DV_G/qinf/V_s/rfd);
% plot([L50(1) L50(1)],[0 0.5],'k:','LineWidth', lineWidth);
% plot([L50(2) L50(2)],[0 0.5],'k:','LineWidth', lineWidth);
% plot([L50(2) max(L)],[0.5 0.5],'k:','LineWidth', lineWidth);
% ylabel('Infection Probability','Interpreter','latex');
% % amend plot
% box on;
% ax = gca; % Get axis to modify
% % ax.XScale = 'log';
% % ax.XLim = [1e5 1e12];
% ax.YAxis(1).Color = 'k';
% ax.YAxis(2).Color = 'k';
% ax.YAxis(1).FontSize = font_size;
% ax.YAxis(2).FontSize = font_size;
% ax.YAxis(1).FontName = font_name;
% ax.YAxis(2).FontName = font_name;
% ax.XAxis.FontSize    = font_size;
% ax.XAxis.FontName    = font_name;
% ax.XLim = [5 12];
% ax.TickLabelInterpreter = 'latex';
% hold off
% 
% set(gcf, 'PaperPositionMode', 'auto');
% print('-depsc',[filename,'.eps']); 
% set(gcf, 'PaperPositionMode', 'manual');
% set(gcf, 'PaperUnits', 'centimeters');
% set(gcf, 'PaperPosition', [0 0 width height]);
% saveas(h_plot,filename,'png');
% close(h_plot);

%% PROBABILITY OF INFECTORS
% width = 6;
% height = 8;
% filename = 'infected_bar';
% h_plot = figure(3);
% N_I   = 0:1:5; % number of infected people
% C     = 1/100; % community infection rate
% N50     = 50; % number of occupants
% P_N50 = exp(N_I*log(C)+(N50-N_I)*log(1-C)+gammaln(N50+1)-gammaln(N_I+1)-gammaln(N50-N_I+1));
% N50     = 5; % number of occupants
% P_N5  = exp(N_I*log(C)+(N50-N_I)*log(1-C)+gammaln(N50+1)-gammaln(N_I+1)-gammaln(N50-N_I+1));
% figure(3);
% p1 = bar(0:1:5,[P_N50',P_N5'],'EdgeColor',[0.9 0.9 0.9]);
% ylabel('Probability, $P(I)$','Interpreter','latex');
% xlabel('Number of Infected People, $I$','Interpreter','latex');
% % amend plot
% box on;
% ax = gca; % Get axis to modify
% ax.YAxis.FontSize = font_size;
% ax.YAxis.FontName = font_name;
% ax.XAxis.FontSize = font_size;
% ax.XAxis.FontName = font_name;
% ax.TickLabelInterpreter = 'latex';
% hold off
% p1(1).FaceColor = [0.3 0.3 0.3];
% p1(2).FaceColor = [0.7 0.7 0.7];
% % set(gcf, 'PaperPositionMode', 'auto');
% % print('-depsc',[filename,'.eps']); 
% set(gcf, 'PaperPositionMode', 'manual');
% set(gcf, 'PaperUnits', 'centimeters');
% set(gcf, 'PaperPosition', [0 0 width height]);
% print('-depsc',[filename,'.eps']); 
% saveas(h_plot,filename,'png');
% close(h_plot);

%% PROPORTION OF PEOPLE INFECTED WITH VIRAL LOAD
% width = 10;
% height = 8;
% filename = 'load_v_infected_pdf';
% h_plot = figure(4);
% % PDF of viral load
% L_mu         = 7;   % mean viral load [log10 RNA copies per ml]
% L_sigma      = 1.4; % stdev of viral load [log10 RNA copies per ml]
% pd           = makedist('Normal','mu',L_mu,'sigma',L_sigma);
% L            = [6:0.1:13];
% L_prob       = pdf(pd,L);
% L            = 10.^L;
% % infection risk
% pop_ratio    = 10;
% V(1)         = 1500; % m3
% V(2)         = V(1)/pop_ratio; % m3
% phi          = 2.36;  % removal rate [1/h]
% T            = 8;     % exposure period [h]
% K            = 0.55;  % lung deposition factor [-]
% k            = 410;   % reciprocal of the probability that a single pathogen initiates an infection [-]
% v            = 1;     % viable fraction [-]
% qsus         = 0.558; % breathing rate [m3/hour]
% qinf         = qsus;  % breathing rate [m3/hour]
% V_s          = 5.05e-13; % Volumetric ratio of exhaled droplets to exhaled air [-]
% DV_G         = K*qsus*T*v/phi; % dose per the product of the volume and emission rate [virions m3.s/RNA copies]
% rfd          = 125e6; % respiratory fluid density [ml/m3]
% G            = qinf*V_s*rfd.*L; % emission rate [RNA copies per hour]
% D50          = (DV_G/V(1)).*G; % dose [virions inhaled]
% P_R1         = 1-exp(-D50./k); % probability of infection
% D5           = DV_G.*G./V(2); % dose [virions inhaled]
% P_R2         = 1-exp(-D5./k); % probability of infection
% % probability of susceptible people
% C = 1/100;
% N50     = 50; % number of occupants
% N5      = 5; % number of occupants
% P_S_50 = (1-C)-(1-C)^N50; % probability of susceptible people present
% P_S_5  = (1-C)-(1-C)^N5;  % probability of susceptible people present
% P_trans_50 = (1-C^N50)-(1-C)^N50; % probability of transmission event
% P_trans_5  = (1-C^N5)-(1-C)^N5;   % probability of transmission event
% I_bar_50 = N50*(C-C^N50)/P_trans_50; % mean number of infected people
% I_bar_5  = N5*(C-C^N5)/P_trans_5;    % mean number of infected people
%
% P_I_50 = 100.*P_R1.*L_prob.*P_S_50.*I_bar_50; % proportion infected
% P_I_5  = 100.*P_R2.*L_prob.*P_S_5.*I_bar_5;   % proportion infected
% x = log10(L); 
% p1 = plot(x,P_I_50,'k','LineWidth',lineWidth);
% hold on;
% plot(x,P_I_5,'--k','LineWidth',lineWidth);
% xlabel({'Viral Load (log$_{10}$ RNA Copies per ml)'},'Interpreter','latex');
% ylabel('Proportion Infected ($\%$)','Interpreter','latex');
% title('');
% % % axis([0,3,0,1]);
% ax = gca; % Get axis to modify
% % ax.XScale = 'log';
% ax.YAxis.FontSize = font_size;
% ax.YAxis.FontName = font_name;
% ax.XAxis.FontSize = font_size;
% ax.XAxis.FontName = font_name;
% ax.TickLabelInterpreter = 'latex';
% 
% set(gcf, 'PaperPositionMode', 'auto');
% print('-depsc',[filename,'.eps']); 
% set(gcf, 'PaperPositionMode', 'manual');
% set(gcf, 'PaperUnits', 'centimeters');
% set(gcf, 'PaperPosition', [0 0 width height]);
% saveas(h_plot,filename,'png');
% close(h_plot);

%% PPI/TR versus ventilation
% number of occupants
% clear all
% filename    = "outputs.xlsx";
% sheets      = {'50','5','summary','PPI'};
% pop_ratio   = 10;
% N(1,1)      = 50; % reference space
% N(2,1)      = N(1)/pop_ratio; % comparator space
% % number of spaces
% spaces(1,1) = 2e5;
% N_pop       = N(1)*spaces(1); % samples
% spaces(2,1) = N_pop/N(2);
% % space volume
% V(1,1)      = 1500; % m3
% V(2,1)      = V(1)/pop_ratio; % m3
% % DOSE TERMS
% rr          = 2.36 - 10*N(1)*3.6/V(1); % removal rate [1/h]
% T           = 8;     % exposure period [h]
% K           = 0.55;  % lung deposition factor [-]
% k           = 410;   % reciprocal of the probability that a single pathogen initiates an infection [-]
% v           = 1;     % viable fraction [-]
% qsus        = 0.558; % breathing rate [m3/hour]
% qinf        = qsus;  % breathing rate [m3/hour]
% C           = 1/100; % community infection rate [-]
% V_s         = 5.05e-13; % Volumetric ratio of exhaled droplets to exhaled air [-]
% rfd         = 125e6; % respiratory fluid density [ml/m3]
% L_mu        = 7;   % mean viral load [log10 RNA copies per ml]
% L_sigma     = 1.4; % stdev of viral load [log10 RNA copies per ml]
% limits{1}   = [0:1:50]; % limits of per capita ventilation rate [l/s/p]
% limits{2}   = 10;       % limits of per capita ventilation rate [l/s/p]
% 
% % CALCULATIONS
% % loop through scenario
% for scen = 1:2
%     counter = 1;
%     PPI = 0;
%     % loop through each space
%     for pcvr = limits{scen}
%         phi  = pcvr*N(scen)*3.6/V(scen)+rr; % per capita ventilation rate [l/s/p]
%         DV_G = K*qsus*T*v/phi; % dose per the product of the volume and emission rate [virions m3.s/RNA copies]
%         for space = 1:spaces(scen)
%             % number of infected people
%             N_I(space,1)   = binornd(N(scen),C);
%             % number of susceptible people
%             N_S(space,1)   = N(scen) - N_I(space);
%             % viral load
%             L(space,1)     = sum(10.^(randn([N_I(space),1]).*L_sigma+L_mu)); % [RNA copies per ml]
%             % emission rate
%             G(space,1)     = qinf*V_s*rfd.*L(space); % [RNA copies per hour]
%             % dose
%             D(space,1)     = DV_G*G(space)/V(scen); % [virions inhaled]
%             % probability of infection
%             P_R(space,1)   = 1-exp(-D(space)/k);
%             % number of susceptible people infected
%             N_S_I(space,1) = binornd(N_S(space),P_R(space));
%             % fraction of susceptible people infected
%             N_S_I_f(space,1) = N_S_I(space)/N_S(space);
%         end
%         % proportion of population infected
%         PPI(counter,1) = sum(N_S_I)/N_pop;
% %         if pcvr == 10 && scen == 1
% %             G_bar   = mean(G);   % mean emission rate
% %             D_bar   = mean(D);   % mean dose
% %             P_R_bar = mean(P_R); % probability of infection
% %             N_S_I_mu_tot = mean(N_S_I); % total mean number of susceptible people infected in each scenario
% %             N_S_I_mu_inf = mean(N_S_I(N_I>0)); % mean number of susceptible people infected in each scenario only when infected people are present
% %             % proportion of population infected
% %             plot_data   = table(N(scen),V(scen),spaces(scen),G_bar,D_bar,P_R_bar,N_S_I_mu_tot,N_S_I_mu_inf,PPI(counter));
% %         end
%         counter = counter + 1;
%     end
%     PPI_out{scen} = PPI;
% end
% % transmission ratio [comparator:reference]
% TR = PPI_out{2}./PPI_out{1};
% 
% writematrix(PPI_out{1},filename,'Sheet',sheets{4});
% 
% plot
% PPI_out{1} = dlmread('vent_v_ppi.dat');
% data{3}    = readtable(filename,'Sheet',sheets{3});
% P_I_5 = data{3}.PPI(2);
% h_plot   = figure(4);
% width    = 10;
% height   = 8;
% filename = 'vent_v_ppi';
% vent_pp = [0:1:50]';
% C=0.55; A=1.9; M=0.0714;
% p1 = plot(vent_pp,C+(A-C).*exp(-M.*vent_pp),'k','LineWidth',lineWidth);
% ylabel({'Big space $PPI$ (\%)'},'Interpreter','latex');
% xlabel('Big space ventilation rate (l/s per person)','Interpreter','latex');
% title('');
% yyaxis right;
% ylabel({'$TR^{-1}$'},'Interpreter','latex');
% ax = gca; % Get axis to modify
% yyaxis left;
% ax.YAxis(1).FontSize = font_size;
% ax.YAxis(1).FontName = font_name;
% ax.YAxis(1).Color = 'k';
% ax.YLim = [0,2.5];                              % CHECK THIS!!!
% ax.YTick = ax.YTick; % forces the retention of ticks when saving
% ax.TickLabelInterpreter = 'latex';
% TR = round(ax.YTick./P_I_5./100,2,'significant');
% yyaxis right;
% ax.YAxis(2).FontSize = font_size;
% ax.YAxis(2).FontName = font_name;
% ax.YAxis(2).Color = 'k';
% ax.YLim = [min(TR) max(TR)];
% ax.YTick = TR;
% ax.TickLabelInterpreter = 'latex';
% ax.YTickLabel = compose('%g',TR);
% 
% % save
% set(gcf, 'PaperPositionMode', 'auto');
% print('-depsc',[filename,'.eps']); 
% set(gcf, 'PaperPositionMode', 'manual');
% set(gcf, 'PaperUnits', 'centimeters');
% set(gcf, 'PaperPosition', [0 0 width height]);
% saveas(h_plot,filename,'png');
% close(h_plot);

%% fraction of people infected
% frac_inf = data{2}.N_S_I;
% % plot
% filename = 'n_infected_hist_5';
% width = 10;
% height = 8;
% h_plot = figure(5);
% h = histogram(frac_inf,'Normalization','probability','FaceColor',[0.7 0.7 0.7]);
% ylabel('Frequency','Interpreter','latex');
% xlabel('Number of People Infected','Interpreter','latex');
% title('');
% ax = gca; % Get axis to modify
% ax.YScale = 'log';
% ax.YAxis.FontSize = font_size;
% ax.YAxis.FontName = font_name;
% ax.XAxis.FontSize = font_size;
% ax.XAxis.FontName = font_name;
% ax.TickLabelInterpreter = 'latex';
% % save
% set(gcf, 'PaperPositionMode', 'auto');
% print('-depsc',[filename,'.eps']); 
% set(gcf, 'PaperPositionMode', 'manual');
% set(gcf, 'PaperUnits', 'centimeters');
% set(gcf, 'PaperPosition', [0 0 width height]);
% saveas(h_plot,filename,'png');
% close(h_plot);

% hc50=histcounts(data{1}.N_S_I,-0.5:1:50.5,'Normalization','probability');
% hc5 =histcounts(data{2}.N_S_I,-0.5:1:5.5,'Normalization','probability');

%% OCCUPANCY V PPI (with 10l/s/p and vol/person preserved)
% Calculation
% number of occupants
% clear all
% filename    = "outputs.xlsx";
% sheets      = {'50','5','summary','PPI'};
% pop_ratio   = 10;
% N(1,1)      = 50; % reference space
% N(2,1)      = N(1)/pop_ratio; % comparator space
% % number of spaces
% spaces(1,1) = 2e5;
% N_pop       = N(1)*spaces(1); % samples
% spaces(2,1) = N_pop/N(2);
% % space volume
% V(1,1)      = 1500; % m3
% V(2,1)      = V(1)/pop_ratio; % m3
% svpp        = V(1)/N(1); % space volume per person
% % Dose terms
% phi         = 2.36;  % removal rate [1/h]
% T           = 8;     % exposure period [h]
% K           = 0.55;  % lung deposition factor [-]
% k           = 410;   % reciprocal of the probability that a single pathogen initiates an infection [-]
% v           = 1;     % viable fraction [-]
% qsus        = 0.558; % breathing rate [m3/hour]
% qinf        = qsus;  % breathing rate [m3/hour]
% C           = 1/100; % community infection rate [-]
% V_s         = 5.05e-13; % Volumetric ratio of exhaled droplets to exhaled air [-]
% DV_G        = K*qsus*T*v/phi; % dose per the product of the volume and emission rate [virions m3.s/RNA copies]
% rfd         = 125e6; % respiratory fluid density [ml/m3]
% L_mu        = 7;   % mean viral load [log10 RNA copies per ml]
% L_sigma     = 1.4; % stdev of viral load [log10 RNA copies per ml]
% occs        = 5:5:150;
% 
% % CALCULATIONS
% % loop through each space
% scen    = 1;
% counter = 1;
% for occ = occs
%     N(scen) = occ;
%     V       = N(scen)*svpp;
%     spaces  = round(N_pop/N(scen),0);
%     for space = 1:spaces
%         % number of infected people
%         N_I(space,1)   = binornd(N(scen),C);
%         % number of susceptible people
%         N_S(space,1)   = N(scen) - N_I(space);
%         % viral load
%         L(space,1)     = sum(10.^(randn([N_I(space),1]).*L_sigma+L_mu)); % [RNA copies per ml]
%         % emission rate
%         G(space,1)     = qinf*V_s*rfd.*L(space); % [RNA copies per hour]
%         % dose
%         D(space,1)     = DV_G*G(space)/V; % [virions inhaled]
%         % probability of infection
%         P_R(space,1)   = 1-exp(-D(space)/k);
%         % number of susceptible people infected
%         N_S_I(space,1) = binornd(N_S(space),P_R(space));
%         % fraction of susceptible people infected
%         N_S_I_f(space,1) = N_S_I(space)/N_S(space);
%     end
%     % proportion of population infected
%     PPI(counter,1) = sum(N_S_I)/N_pop;
%     counter = counter + 1;
%     clear N_S_I N_S_I P_R D G L N_S N_I
% end
% transmission ratio [comparator:reference]
% TR = PPI_out{1}(1)./PPI_out{1};
% 
% % save and retrieve
% filename = 'occ_v_PPI';
% dlmwrite([filename,'.dat'],[occs',PPI]);

% 
% plot
% filename = 'occ_v_PPI';
% plot_data = dlmread([filename,'.dat']);
% PPI  = plot_data(:,2);
% occs = plot_data(:,1)';
% P_I_5    = PPI(1);
% h_plot   = figure(5);
% width    = 10;
% height   = 8;
% 
% m = 0.0176; % see data spreadsheet
% c = 1.95;
% 
% p1 = plot([0,1,occs]',c.*(1-exp(-[0,1,occs]'.*m)),'k','LineWidth',lineWidth);
% ylabel({'Big space $PPI$ (\%)'},'Interpreter','latex');
% xlabel('Big space $N$','Interpreter','latex');
% title('');
% yyaxis right;
% ylabel({'$TR^{-1}$'},'Interpreter','latex');
% ax = gca; % Get axis to modify
% yyaxis left;
% ax.YAxis(1).FontSize = font_size;
% ax.YAxis(1).FontName = font_name;
% ax.YAxis(1).Color = 'k';
% ax.YLim = [0,2];                              % CHECK THIS!!!
% ax.YTick = ax.YTick; % forces the retention of ticks when saving
% ax.TickLabelInterpreter = 'latex';
% TR = round(ax.YTick./P_I_5./100,1);
% yyaxis right;
% ax.YAxis(2).FontSize = font_size;
% ax.YAxis(2).FontName = font_name;
% ax.YAxis(2).Color = 'k';
% ax.YLim = [min(TR) max(TR)];
% ax.YTick = TR;
% ax.TickLabelInterpreter = 'latex';
% ax.YTickLabel = compose('%g',TR);
% ax.XTick = 0:25:150;
% 
% % save
% set(gcf, 'PaperPositionMode', 'auto');
% print('-depsc',[filename,'.eps']); 
% set(gcf, 'PaperPositionMode', 'manual');
% set(gcf, 'PaperUnits', 'centimeters');
% set(gcf, 'PaperPosition', [0 0 width height]);
% saveas(h_plot,filename,'png');
% close(h_plot);

%% OCCUPANCY V PPI (V=1500m3 and  ventilation rate is 500l/s)
% % Calculation
% % number of occupants
% clear all
% filename    = "outputs.xlsx";
% sheets      = {'50','5','summary','PPI'};
% N_pop       = 50*2e5; % % number of spaces/samples
% V           = 1500;  % space volume [m3]
% phi         = 2.36;  % removal rate [1/h]
% T           = 8;     % exposure period [h]
% K           = 0.55;  % lung deposition factor [-]
% k           = 410;   % reciprocal of the probability that a single pathogen initiates an infection [-]
% v           = 1;     % viable fraction [-]
% qsus        = 0.558; % breathing rate [m3/hour]
% qinf        = qsus;  % breathing rate [m3/hour]
% C           = 1/100; % community infection rate [-]
% V_s         = 5.05e-13; % Volumetric ratio of exhaled droplets to exhaled air [-]
% D_G         = K*qsus*T*v/phi/V; % dose per the product of the volume and emission rate [virions m3.s/RNA copies]
% rfd         = 125e6; % respiratory fluid density [ml/m3]
% L_mu        = 7;   % mean viral load [log10 RNA copies per ml]
% L_sigma     = 1.4; % stdev of viral load [log10 RNA copies per ml]
% occs        = [5,50,75,100,125,150];
% 
% % CALCULATIONS
% % loop through each space
% scen    = 1;
% counter = 1;
% for occ = occs
%     N(scen) = occ;
%     spaces  = round(N_pop/N(scen),0);
%     for space = 1:spaces
%         % number of infected people
%         N_I(space,1)   = binornd(N(scen),C);
%         % number of susceptible people
%         N_S(space,1)   = N(scen) - N_I(space);
%         % viral load
%         L(space,1)     = sum(10.^(randn([N_I(space),1]).*L_sigma+L_mu)); % [RNA copies per ml]
%         % emission rate
%         G(space,1)     = qinf*V_s*rfd.*L(space); % [RNA copies per hour]
%         % dose
%         D(space,1)     = D_G*G(space); % [virions inhaled]
%         % probability of infection
%         P_R(space,1)   = 1-exp(-D(space)/k);
%         % number of susceptible people infected
%         N_S_I(space,1) = binornd(N_S(space),P_R(space));
%         % fraction of susceptible people infected
%         N_S_I_f(space,1) = N_S_I(space)/N_S(space);
%     end
%     % proportion of population infected
%     PPI(counter,1) = sum(N_S_I)/N_pop;
%     counter = counter + 1;
%     clear N_S_I N_S_I P_R D G L N_S N_I
% end
% 
% % plot
% % ppi_data = dlmread([filename,'.dat']);
% data{3}  = readtable(filename,'Sheet',sheets{3});
% filename = 'occ_v_PPI_2';
% occs     = [5,50,75,100,125,150];
% P_I_5    = data{3}.PPI(2);
% h_plot   = figure(6);
% width    = 10;
% height   = 8;
% 
% m = 0.025; % see data spreadsheet
% c = -0.0103;
% plot([0,occs]',(c + [0,occs]'.*m),'k','LineWidth',lineWidth);
% hold on;
% tr1 = (P_I_5*100-c)/m;
% plot([0,150],[P_I_5*100,P_I_5*100],'k:','LineWidth',lineWidth);
% plot([tr1,tr1],[0,P_I_5*100],'k:','LineWidth',lineWidth);
% ylabel({'Big space $PPI$ (\%)'},'Interpreter','latex');
% xlabel('Big space $N$','Interpreter','latex');
% title('');
% yyaxis right;
% ylabel({'$TR^{-1}$'},'Interpreter','latex');
% ax = gca; % Get axis to modify
% yyaxis left;
% ax.YAxis(1).FontSize = font_size;
% ax.YAxis(1).FontName = font_name;
% ax.YAxis(1).Color = 'k';
% ax.YLim = [0,4];                              % CHECK THIS!!!
% ax.YTick = ax.YTick; % forces the retention of ticks when saving
% ax.TickLabelInterpreter = 'latex';
% TR = round(ax.YTick./P_I_5./100,1);
% yyaxis right;
% ax.YAxis(2).FontSize = font_size;
% ax.YAxis(2).FontName = font_name;
% ax.YAxis(2).Color = 'k';
% ax.YLim = [min(TR) max(TR)];
% ax.YTick = TR;
% ax.TickLabelInterpreter = 'latex';
% ax.YTickLabel = compose('%g',TR);
% ax.XTick = 0:25:150;
% 
% % save
% set(gcf, 'PaperPositionMode', 'auto');
% print('-depsc',[filename,'.eps']); 
% set(gcf, 'PaperPositionMode', 'manual');
% set(gcf, 'PaperUnits', 'centimeters');
% set(gcf, 'PaperPosition', [0 0 width height]);
% saveas(h_plot,filename,'png');
% close(h_plot);

%% Community Infection Rate on PPI
% % number of occupants
% pop_ratio   = 10;
% N(1,1)      = 50; % reference space
% N(2,1)      = N(1)/pop_ratio; % comparator space
% % number of spaces
% spaces(1,1) = 2e5;
% N_pop       = N(1)*spaces(1); % samples
% spaces(2,1) = N_pop/N(2);
% % space volume
% V(1,1)      = 1500; % m3
% V(2,1)      = V(1)/pop_ratio; % m3
% % DOSE TERMS
% phi         = 2.36;  % removal rate [1/h]
% T           = 8;     % exposure period [h]
% K           = 0.55;  % lung deposition factor [-]
% k           = 410;   % reciprocal of the probability that a single pathogen initiates an infection [-]
% v           = 1;     % viable fraction [-]
% qsus        = 0.558; % breathing rate [m3/hour]
% qinf        = qsus;  % breathing rate [m3/hour]
% C           = 1/100; % community infection rate [-]
% V_s         = 5.05e-13; % Volumetric ratio of exhaled droplets to exhaled air [-]
% DV_G        = K*qsus*T*v/phi; % dose per the product of the volume and emission rate [virions m3.s/RNA copies]
% rfd         = 125e6; % respiratory fluid density [ml/m3]
% L_mu        = 7;   % mean viral load [log10 RNA copies per ml]
% L_sigma     = 1.4; % stdev of viral load [log10 RNA copies per ml]
% iCIR        = [1:1:9,10:10:100,200,300,600,1000]; % community infection rate [-]
% CIR         = 1./iCIR;
% 
% % loop through scenario
% for scen = 1:2
%     counter = 1;
%     for C = CIR
%         % loop through each space
%         for space = 1:spaces(scen)
%             % number of infected people
%             N_I(space,1)   = binornd(N(scen),C);
%             % number of susceptible people
%             N_S(space,1)   = N(scen) - N_I(space);
%             % viral load
%             L(space,1)     = sum(10.^(randn([N_I(space),1]).*L_sigma+L_mu)); % [RNA copies per ml]
%             % emission rate
%             G(space,1)     = qinf*V_s*rfd.*L(space); % [RNA copies per hour]
%             % dose
%             D(space,1)     = DV_G*G(space)/V(scen); % [virions inhaled]
%             % probability of infection
%             P_R(space,1)   = 1-exp(-D(space)/k);
%             % number of susceptible people infected
%             N_S_I(space,1) = binornd(N_S(space),P_R(space));
%             % fraction of susceptible people infected
%             N_S_I_f(space,1) = N_S_I(space)/N_S(space);
%         end
%         PPI{scen}(counter,1)   = sum(N_S_I)/N_pop; % proportion of population infected
%         counter = counter + 1;
%         clear N_S_I N_S_I P_R D G L N_S N_I
%     end
% end
% 
% % plot
% filename = 'CIR_v_PPI';
% % ppi_data = dlmread([filename,'.dat']);
% iCIR     = [2:0.1:9.9,10:10:1000]; % community infection rate [-]
% CIR      = 1./iCIR;
% % P_I_5    = PPI(1);
% P_I_5    = data{3}.PPI(2);
% h_plot   = figure(6);
% width    = 10;
% height   = 8;
% 
% c50 = 0.0019;
% m50 = 1.0714;
% c5  = 0.0004103;
% m5  = 0.3409;
% plot([1,iCIR],[0,(82.5.*CIR.^3-198.74.*CIR.^2+125.82.*CIR)],'k','LineWidth',lineWidth);
% hold on;
% plot([1,iCIR],[0,(4.76.*CIR.^3-43.31.*CIR.^2+38.19.*CIR)],'--k','LineWidth',lineWidth);
% ylabel({'Big space $PPI$ (\%)'},'Interpreter','latex');
% xlabel('Community infection rate, $C^{-1}$','Interpreter','latex');
% title('');
% yyaxis right;
% plot([1,max(iCIR)],[3.16,3.16],'-.k','LineWidth',lineWidth);
% ylabel({'$TR^{-1}$'},'Interpreter','latex');
% ax = gca; % Get axis to modify
% yyaxis left;
% ax.YAxis(1).FontSize = font_size;
% ax.YAxis(1).FontName = font_name;
% ax.YAxis(1).Color = 'k';
% ax.YLim = [0,3];                              % CHECK THIS!!!
% ax.YTick = ax.YTick; % forces the retention of ticks when saving
% ax.TickLabelInterpreter = 'latex';
% TR = round(ax.YTick./2,1);
% yyaxis right;
% ax.YAxis(2).FontSize = font_size;
% ax.YAxis(2).FontName = font_name;
% ax.YAxis(2).Color = 'k';
% ax.YLim = [0 6];
% % ax.YTick = TR;
% ax.TickLabelInterpreter = 'latex';
% % ax.YTickLabel = compose('%g',TR);
% ax.XLim= [0,500];
% 
% % save
% set(gcf, 'PaperPositionMode', 'auto');
% print('-depsc',[filename,'.eps']); 
% set(gcf, 'PaperPositionMode', 'manual');
% set(gcf, 'PaperUnits', 'centimeters');
% set(gcf, 'PaperPosition', [0 0 width height]);
% saveas(h_plot,filename,'png');
% close(h_plot);


%% fraction of people infected
% data{1} = readtable(filename,'Sheet',sheets{1});
% frac_inf = data{1}.N_S_I;
% % plot
% filename = 'n_infected_hist';
% width = 10;
% height = 8;
% h_plot = figure(7);
% histogram(frac_inf,'Normalization','probability','FaceColor',[0.7 0.7 0.7]);
% ylabel('Frequency','Interpreter','latex');
% xlabel('Number of People Infected','Interpreter','latex');
% title('');
% ax = gca; % Get axis to modify
% ax.YScale = 'log';
% ax.YAxis.FontSize = font_size;
% ax.YAxis.FontName = font_name;
% ax.XAxis.FontSize = font_size;
% ax.XAxis.FontName = font_name;
% ax.TickLabelInterpreter = 'latex';
% % save
% set(gcf, 'PaperPositionMode', 'auto');
% print('-depsc',[filename,'.eps']); 
% set(gcf, 'PaperPositionMode', 'manual');
% set(gcf, 'PaperUnits', 'centimeters');
% set(gcf, 'PaperPosition', [0 0 width height]);
% saveas(h_plot,filename,'png');
% close(h_plot);
```
